# Nonstructural Protein 1 (NS1)-based IgG FcγR competition Enzyme-Linked Immunosorbent Assays (ELISAs) for highly specific serological profiling of previous flavivirus exposures

**DOI:** 10.64898/2026.11.17.26365888

**Authors:** Christa Ehmen, Rutineia Ferraz, Jörg Blessmann, Vatsana Sopraseuth, Latdamone Keoviengkhone, Ana Bispo, Flavia Levy, Iryna Demchyshyna, Anna Papa, Mateja Jelovsek, Misa Korva, Lars Matthies, Michael Ramharter, Tamara Nordmann, Martin Gabriel, Stephan Günther, Simone Kann, Ronald von Possel, Yang Liu, Herbert Schmitz, Jonas Schmidt-Chanasit, Petra Emmerich, Christina Deschermeier

**Author notes:** Corresponding author: Christina Deschermeier Panadea Diagnostics GmbH Bernhard-Nocht-Str. 74 20259 Hamburg. **List of human genes:** *Hs*FcµR: symbol: FCMR; name: Fc fragment of IgM receptor; other symbols and names/synonyms: FAIM3/TOSO *Hs*FcγRIIA: symbol: FCGR2A; name: Fc fragment of IgG receptor IIa; other symbols and names/synonyms: CD32A.

## Abstract

**Background:** Due to extensive antibody cross-reactivity, serological discrimination of IgG antibodies elicited by exposure to different flaviviruses remains challenging. We aimed to develop and validate highly discriminative IgG ELISAs employing an Fcγ receptor (FcγR) capture platform and recombinant competitor molecules to suppress cross-reactive responses.

**Methods:** IgG ELISAs targeting Dengue virus (DENV), Zika virus (ZIKV), and West Nile virus (WNV) were established using immobilized human FcγRIIA for antibody capture and soluble, labeled nonstructural protein 1 (NS1) as antigen. To suppress flavivirus cross-reactivity, target-specific competitor molecule formulations (“Specificity Enhancers” (*SEs*)), comprising unlabeled heterologous NS1 proteins, were incorporated. ELISAs were validated using comprehensive serum panels from Europe, Asia, Africa, and the Americas.

**Results:** Validation revealed reliable detection of seroconversion following DENV, ZIKV, and WNV infections, as well as DENV vaccination. In sera from flavivirus-endemic regions, substantial cross-reactivity was observed in the absence of *SEs*, predominantly in samples showing strong reactivity to the corresponding homologous antigen. Addition of *SEs* significantly improved specificity without compromising sensitivity in most sample panels. In complex exposure settings, analysis of differential signal modulation by individual *SE* components may provide insights into possible exposure histories. Notably, serological discrimination between IgG antibodies elicited by exposure to different WNV lineages could be achieved by combining lineage-homologous NS1 antigens with lineage-heterologous competitor molecules.

**Conclusions:** The flavivirus (NS1) IgG FcγR *SE* ELISA platform enables enhanced differentiation of flavivirus-specific IgG responses in epidemiologically complex settings. Its modular design allows versatile adaptation to newly emerging flaviviruses and supports high-resolution seroepidemiological investigations in One Health contexts.

**Importance:** Cross-reactivity of anti-flavivirus IgG in serological assays is a major obstacle in flavivirus serology, hampering both acute diagnostics and seroprevalence studies. By combining Fcγ-receptor (FcγR)-mediated antibody capture with target-specific competitor molecules, we developed and validated enzyme-linked immunosorbent assays (ELISAs) for the highly discriminative detection of anti-Dengue virus (DENV), anti-Zika virus (ZIKV), and anti-West Nile virus (WNV) nonstructural protein 1 (NS1) IgG antibodies in human sera. The novel assays enable seroprevalence estimates that are not confounded by co-circulating flaviviruses or flavivirus vaccinations. Due to the modular design, this serological platform technology can be flexibly extended to further flaviviruses as they emerge.

## INTRODUCTION

The *Flaviviridae* family comprises a large group of enveloped, positive-sense, single-stranded RNA viruses of major global public health relevance. Several arthropod-borne, human-pathogenic members of the *Orthoflavivirus* genus co-circulate in many regions worldwide, including Africa, the Americas, Europe, and Asia, notably Dengue virus (DENV), Zika virus (ZIKV), West Nile virus (WNV), Yellow fever virus (YFV), Japanese encephalitis virus (JEV), and Tick-borne encephalitis virus (TBEV) (1). Vaccines against JEV, TBEV, YFV, and, more recently, DENV are widely used in endemic areas and among travelers, resulting in substantial levels of flavivirus immunity in certain populations.

Humoral immune responses to flaviviruses are highly complex and heterogeneous. The magnitude and specificity of antibody responses are influenced by multiple factors, including the infecting virus species, vaccination history, and the order and temporal spacing of sequential flavivirus exposures (2–6). Due to the high sequence similarities between the different flaviviruses, cross-reactive antibodies are frequently induced or boosted upon secondary exposure (7). This extensive serological cross-reactivity substantially hampers reliable determination of flavivirus-specific seroprevalence and may lead to erroneous epidemiological conclusions, particularly when the highly conserved envelope protein (EP) is used as antigen in antibody binding assays (8). Point mutations in the EP fusion loop domain, a predominant target of cross-reacting antibodies, have been shown to significantly reduce but not completely eliminate cross-reactivity (8–12). In recent years, nonstructural protein 1 (NS1) has emerged as an alternative antigen exhibiting generally improved specificity (13–18). Reduced cross-reactivity of NS1-based assays has been attributed to structural rather than purely sequence-based differences: although predicted NS1 B-cell epitopes show high sequence identity to corresponding regions in other flaviviruses, variations in non-conserved epitope-flanking residues alter electrostatic surface potentials, potentially limiting heterologous antibody binding (19). Nevertheless, the use of NS1 as antigen does not fully eliminate serological cross-reactivity (16, 20, 21), underscoring the persistent diagnostic challenges in flavivirus serology.

Recently, we have shown that highly discriminative flavivirus serological testing can be achieved by combining a patented IgG Fcγ receptor-based ELISA technology (22, 23) with the use of EP domain III (EDIII) as antigen and recombinant competitor molecules for efficient suppression of cross-reactive responses (20, 24). In this study, we applied this technological concept to develop ELISAs targeting NS1-specific IgG antibodies against DENV, ZIKV, and WNV. We evaluated the discriminatory potential of these assays using serum samples from flavivirus-endemic regions from Europe, the Americas, Asia, and Africa, demonstrating their applicability for improved serological differentiation in complex epidemiological settings.

## MATERIALS AND METHODS

### Recombinant flavivirus NS1 proteins

DENV1-4 and JEV NS1 proteins produced in HEK293T cells were purchased from The Native Antigen Company (Oxford, UK). Amplicons encoding ZIKV, WNV lineage 1, WNV lineage 2, USUV, and YFV NS1 proteins were generated by PCR using either random-primed cDNA transcribed from virus RNA or synthesized cDNA (codon-optimized for expression in mammalian cells; ThermoFisher Scientific, Waltham, USA) and inserted by in-fusion cloning (TaKaRa, Kusatsu, Japan) into pSecTag2-A (ThermoFisher Scientific, Waltham, USA) cut with HindIII/XhoI. Insert sequences were verified using Sanger sequencing (Microsynth Seqlab GmbH, Göttingen, Germany). Plasmids were transiently transfected into HEK293T cells with polyethylenimine. 72 hours post transfection, cell culture supernatant was collected and cleared by centrifugation. Recombinant, His_6_-tagged NS1 proteins were purified using Ni-nitrilotriacetic acid (NTA) chromatography under native conditions followed by size-exclusion chromatography (SEC) on a Superdex 200 10/300 column (GE Healthcare, Chicago, USA). Proteins were stored at-80°C until use.

### Horseradish peroxidase (HRP) labeling of recombinant NS1 proteins

Recombinant NS1 proteins were covalently coupled to sodium periodate-activated HRP as previously described (25, 26). Labeled proteins were stored at-20°C until use.

### Human sera

Specimens accompanied by the corresponding anamnestic and pre-characterization information were provided by multiple institutions (**Supplemental Table 1**). Data privacy protection was guaranteed by anonymization of samples. Where required, ethical approval was obtained from the appropriate authorities, including the Ethics Committee of the Oswaldo Cruz Foundation/Brazil (under CAAE 90249218.6.1001.5248, approval no. 6.824.149), the Ethics Committee of the Hospital Rosario Pumarejo de Lopez of Valledupar/Colombia, the Ethics Committee of Lao People’s Democratic Republic (no. 030/NECHR), the National Medical Ethics Committee of Republic of Slovenia (no. 131/06/13, no. 152/06/13), and the Medical Association Hamburg/Germany (no. PV4608, no. 2020-10162-BO-ff, no. 2023-101107-BO).

### Rabbit hyperimmune sera

Rabbits were immunized with purified recombinant proteins at Eurogentec (Seraing, Belgium).

### Flavivirus IgG Indirect Immunofluorescence Testing (IIFT)

Immunofluorescence analysis using flavivirus infected Vero cells was performed as described previously (27).

### DENV (NS1) IgM FcµR ELISA, ZIKV (NS1) IgM FcµR ELISA

Assays were performed as described previously (26). Briefly, serum samples were co-incubated at a final dilution of 1:100 together with an HRP-labeled recombinant NS1 antigen in a 96-well plate coated with recombinant *Hs*FcµR. Following a washing step, the bound IgM/antigen immune complexes were visualized by application of the colorimetric HRP-substrate tetramethylbenzidine (TMB). After stopping the enzymatic reaction, the assay result was generated by measuring the absorbance (*A*) of the solution in the well at 450/620 nm.

### Flavivirus (NS1) IgG FcγR ELISAs

Assay protocols were established on the FcγR platform as described previously for other target pathogens/flavivirus antigens (24–26, 28). Briefly, serum samples were co-incubated at a final dilution of 1:100 together with an HRP-labeled recombinant NS1 antigen (**Table 1**) and, if stated, a Specificity Enhancer (*SE*) reagent comprising non-labeled NS1 proteins from other flaviviruses (**Table 1**) in a 96-well plate coated with recombinant *Hs*FcγRIIA. Following a washing step, the bound IgG/antigen immune complexes were visualized by application of the colorimetric HRP-substrate tetramethylbenzidine (TMB). After stopping the enzymatic reaction, the assay result was generated by measuring the absorbance (*A*) of the solution in the well at 450/620 nm. Results were either reported as raw data (*A*450 – *A*620) or index values (raw data normalized to assay cut-off).

**Table 1.** Flavivirus (NS1) IgG FcyR ELISAs: antigens and target-specific Specificity Enhancer (*SE)* reagents. DENV: Dengue virus, ZIKV: Zika virus, WNV: West Nile virus, USUV: Usutu virus, YFV: Yellow fever virus, lin: lineage, NS1: nonstructural protein 1

| **IgG FcγR ELISA** | **Labeled antigen** | **Accession no. antigen** | **Specificity Enhancer (*SE*)** |
|---|---|---|---|
| DENV (NS1) | DENV-1/2/3/4 NS1 | AAB70694.1, AAB58782.1, AAM51537.1, AAK01233.1 | none |
| DENV (NS1) *SE* |  |  | ZIKV/WNV/JEV/USUV/YFV NS1 |
| ZIKV (NS1) | ZIKV NS1 | AYU74865.1 | none |
| ZIKV (NS1) *SE* |  |  | DENV1-4/WNV/JEV/USUV/YFV NS1 |
| WNV lin1 (NS1) | WNV lin1 NS1 | QXU69896.1 | none |
| WNV lin1 (NS1) *SE* |  |  | DENV1-4/ZIKV/JEV/USUV/YFV NS1 |
| WNV lin2 (NS1) | WNV lin2 NS1 | AZK36003.1 | none |
| WNV lin2 (NS1) *SE* |  |  | DENV1-4/ZIKV/JEV/USUV/YFV NS1 |

### Other commercially available flavivirus IgG ELISAs

The Panbio Dengue IgG Indirect ELISA (Abbott Laboratories, Abbott Park, USA) and the West Nile Virus IgG DxSelect ELISA (Focus Diagnostics, Cypress, USA) were used according to the manufacturers’ instructions.

### Statistical analysis

Calculation of 95% confidence intervals (CIs) according to the modified Wald method was done using GraphPad QuickCalcs. Statistical testing (paired t test) was performed using GraphPad Prism (version 10.4.1). Receiver operating characteristics (ROC) analyses were performed using Medcalc (version 24.0.1).

## RESULTS

### Development of DENV, ZIKV, and WNV (NS1) IgG FcγR ELISAs

As shown previously, combination of recombinant *Hs*FcγRIIA as immobilized capture molecule with HRP-labeled recombinant antigens and unlabeled competitor molecules allows the development of ELISAs optimized for differential detection of anti-flavivirus EDIII IgG antibodies (20, 24). Leveraging this technological concept, prototypic IgG FcγR ELISAs targeting three medically relevant flaviviruses (DENV, ZIKV, WNV) employing HRP-labeled, recombinant NS1 proteins as antigens were developed (**Table 1**). For suppression of false positive signals arising from cross-reactive antibodies elicited by other human pathogenic flaviviruses than the respective pathogen of interest, a target-specific mix of recombinant, unlabeled NS1 proteins, designated as “Specificity Enhancer (*SE*)” was optionally added (**Table 1).** Antigen and *SE* concentrations were titrated (data not shown) to achieve minimized assay background, efficient use of the dynamic range of detection (**Supplemental Figure 1A**) for true positives, and optimal suppression of signals elicited by cross-reactive antibodies. Assay cut-offs were defined based on receiver operating characteristic (ROC) analyses (**Supplemental Figure 1B-E**).

### Reproducibility

All assays (DENV (NS1) IgG FcγR, ZIKV (NS1) IgG FcγR, WNV lin1/2 (NS1) IgG FcγR, each without (w/o) and with (w/) target-specific *SEs*) were highly reproducible with mean intra-assay coefficients of variation (CVs) ≤ 5% and mean inter-assay CVs of ≤ 10% for positive signals (**Tables 2A-D**).

### Specificity (complete panel)

Assay specificity was assessed by analyzing serum samples from healthy donors with and without previous flavivirus vaccinations, serum samples from patients with confirmed flavivirus infections (collected both from European travelers and in endemic countries), as well as samples from patients with infections caused by non-flavivirus pathogens such as CHIKV, SARS-CoV-2 and *Plasmodium* (**Supplemental Table 1, Table 2A-D**). Overall assay specificities were found to be 96.9% (595/614) and 98.0% (602/614) for the DENV (NS1) IgG FcγR ELISA w/o and w/ *SE*, 90.6% (604/667) and 100.0% (667/667) for the ZIKV (NS1) IgG FcγR ELISA w/o and w/ *SE*, 91.9% (485/528) and 99.1% (523/528) for the WNV lin1 (NS1) IgG FcγR ELISA w/o and w/ *SE*, and 93.6% (508/543) and 99.6% (541/543) for the WNV lin2 (NS1) IgG FcγR ELISA w/o and w/ *SE*, respectively (**Table 2A-D**).

**Table 2.** A. DENV (NS1) IgG FcyR ELISA validation data. ^1^IgG seropositivity/-conversion detected in 12/12 patients, ^2^samples testing negative for DENV NS1 IgG are from early acute phase (4/4 DENV NS1 IgM positive, 3/4 PCR and/or NS1 antigen positive), ^3^1/8 sample equivocal (sample is DENV NS1 IgM positive), ^4^1 sample testing negative in DENV ELISA w/ *SE* strongly reactive with JEV NS1, ^5^2/4 samples testing negative in the ELISA also tested negative in DENV IgG IIFT, 4/4 samples do not react with DENV1-4 EDIII, ^6^2 samples testing positive in ELISA also tested positive in flavivirus IgG IIFT (DENV IgG titres 1:1280 and 1:640, respectively), ^7^2 samples testing positive in ELISA tested negative in flavivirus IgG IIFT, ^8^1 sample testing positive in ELISA tested also strongly positive in flavivirus IgG IIFT (WNV IgG titre 1:10240, DENV IgG titre 1:5120), ^9^for the 7 samples testing positive in ELISA, presence of anti-flavivirus IgG was confirmed by independent serological testing (**Supplemental** Table 2). Abbreviations: w/o: without, w/: with, DENV: Dengue virus, ZIKV: Zika virus, WNV: West Nile virus, YF: Yellow fever, TBEV: Tick-borne encephalitis virus, CHIKV: Chikungunya virus, CV: coefficient of variation, CI: confidence interval, SARS-CoV-2: Severe Acute Respiratory Syndrome Corona virus 2, WHO: World Health Organization, N: number of sera or patients tested.

| **DENV (NS1) IgG FcγR ELISA** |  |  | **w/o *SE*** | **w/ *SE*** |
|---|---|---|---|---|
| **Assay reagents** | HRP-labeled antigen |  | DENV1/2/3/4 NS1 | DENV1/2/3/4 NS1 |
|  | Specificity Enhancer (*SE*) |  | none | ZIKV/WNV/JEV/USUV/YFV NS1 |
| **Reproducibility** | Intra-assay CV |  | ≤ 5% | ≤ 5% |
|  | Inter-assay CV |  | ≤ 10% | ≤ 10% |
| **Sensitivity** | **Serum panel** | **N** | **n positive, % sensitivity (95% CI)** | **n positive, % sensitivity (95% CI)** |
|  | DENV infection (German travelers, longitudinal), panel A | 12 | 12, 100.0 (71.8 – 100.0) | 12, 100.0 (71.8 – 100.0)<sup>1</sup> |
|  | DENV infection (German travelers), panel B | 11 | 8, 72.7 (42.9 – 90.8) | 7, 63.6 (35.2 – 85.0)<sup>2</sup> |
|  | DENV infection (German travelers, residual samples) | 8 | 7, 87.5 (50.8 – 99.9) | 7, 87.5 (50.8 – 99.9)<sup>3</sup> |
|  | DENV infection (Lao PDR residents), S1 | 35 | 23, 65.7 (49.1 – 79.2) | 23, 65.7 (49.1 – 79.2) |
|  | DENV infection (Lao PDR residents), S2 | 35 | 35, 100.0 (88.2 – 100.0) | 34, 97.1 (84.2 – 100.0)<sup>4</sup> |
|  | DENV infection (Colombia residents), S1 | 12 | 9, 75.0 (46.1 – 91.7) | 9, 75.0 (46.1 – 91.7) |
|  | DENV infection (Colombia residents), S2 | 12 | 12, 100.0 (71.8 – 100.0) | 12, 100.0 (71.8 – 100.0) |
|  | DENV naïve vaccinees (Germany residents), S3 | 24 | 24, 100.0 (83.7 – 100.0) | 24, 100.0 (83.7 – 100.0) |
|  | DENV preimmune vaccinees (Germany residents), S0 | 25 | 21, 84.0 (64.7 – 94.2) | 21, 84.0 (64.7 – 94.2)<sup>5</sup> |
| **Specificity** | **Serum panel** | **N** | **n negative, % specificity (95% CI)** | **n negative, % specificity (95% CI)** |
|  | Healthy donors (Germany residents) | 42 | 40, 95.2 (83.3 – 99.5) | 40, 95.2 (83.3 – 99.5)<sup>6</sup> |
|  | DENV naïve vaccinees (Germany residents), S0 (baseline) | 24 | 24, 100.0 (83.7 – 100.0) | 24, 100.0 (83.7 – 100.0) |
|  | Healthy donors (Ukraine residents) | 354 | 351, 99.2 (97.4 – 99.8) | 352, 99.4 (97.8 – 99.9)<sup>7</sup> |
|  | ZIKV infection (German travelers) | 9 | 8, 88.9 (54.3 – 100.0) | 9, 100.0 (65.5 – 100.0) |
|  | WNV infection (Canada residents) | 18 | 15, 83.3 (60.0 – 95.0) | 17, 94.4 (72.3 – 100.0)<sup>8</sup> |
|  | YF vaccinees (German travelers) | 91 | 90, 98.9 (93.4 – 100.0) | 91, 100.0 (95.1 – 100.0) |
|  | TBEV NS1 IgG positive | 2 | 2, 100.0 (29.0 – 100.0) | 2, 100.0 (29.0 – 100.0) |
|  | CHIKV infection (WHO IgG standard, 1502/19) | 1 | 1, 100.0 (16.7 – 100.0) | 1, 100.0 (16.7 – 100.0) |
|  | SARS-CoV-2 infection (Germany residents) | 19 | 19, 100.0 (80.2 – 100.0) | 19, 100.0 (80.2 – 100.0) |
|  | *Plasmodium* infection (residual samples) | 54 | 45, 83.3 (71.0 – 91.2) | 47, 87.0 (75.3 – 93.9)<sup>9</sup> |
|  | Complete panel | 614 | 595, 96.9 (95.2 – 98.0) | 602, 98.0 (96.6 – 98.9) |

**Table 2B.** ZIKV (NS1) IgG FcyR ELISA validation data. ^1^IgG seroconversion detected in 8/8 patients, ^2^patients testing ZIKV (NS1) IgG ELISA negative were found to be ZIKV (NS1) IgM ELISA positive. Abbreviations: w/o: without, w/: with, DENV: Dengue virus, ZIKV: Zika virus, WNV: West Nile virus, YF: Yellow fever, TBEV: Tick-borne encephalitis virus, CHIKV: Chikungunya virus, CV: coefficient of variation, CI: confidence interval, WHO: World Health Organization, N: number of sera or patients tested.

| **ZIKV (NS1) IgG FcγR ELISA** |  |  | **w/o *SE*** | **w/ *SE*** |
|---|---|---|---|---|
| **Assay reagents** | HRP-labeled antigen |  | ZIKV NS1 | ZIKV NS1 |
|  | Specificity Enhancer (*SE*) |  | none | DENV1-4/WNV/JEV/USUV/YFV NS1 |
| **Reproducibility** | Intra-assay CV |  | ≤ 5% | ≤ 5% |
|  | Inter-assay CV |  | ≤ 10% | ≤ 10% |
| **Sensitivity** | **Serum panel** | **N** | **n positive, % sensitivity (95% CI)** | **n positive, % sensitivity (95% CI)** |
|  | ZIKV infection (German travelers, longitudinal) | 8 | 8, 100.0 (62.8 – 100.0) | 8, 100.0 (62.8 – 100.0)<sup>1</sup> |
|  | ZIKV infection (German travelers, acute) | 9 | 5, 50.0 (23.7 – 76.3) | 5, 50.0 (23.7 – 76.3)<sup>2</sup> |
|  | ZIKV PRNT pos (Brazil residents, adults) | 9 | 8, 88.9 (54.3 – 100.0) | 7, 77.8 (44.3 – 94.7) |
|  | ZIKV PRNT pos (Brazil residents, children) | 5 | 5, 100.0 (51.1 – 100.0) | 5, 100.0 (51.1 – 100.0) |
|  | ZIKV infection (WHO IgG standard, 16/352) | 1 | 1, 100.0 (16.7 – 100.0) | 1, 100.0 (16.7 – 100.0) |
| **Specificity** | **Serum panel** | **N** | **n negative, % specificity (95% CI)** | **n negative, % specificity (95% CI)** |
|  | Healthy donors (Germany residents) | 42 | 42, 100.0 (90.0 – 100.0) | 42, 100.0 (90.0 – 100.0) |
|  | Healthy donors (Ukraine residents) | 200 | 200, 100.0 (97.7 – 100.0) | 200, 100.0 (97.7 – 100.0) |
|  | Healthy donors (Lao PDR residents) | 20 | 17, 85.0 (63.1 – 95.6) | 20, 100.0 (81.0 – 100.0) |
|  | Healthy donors (Colombia residents) | 134 | 107, 79.9 (72.2 – 85.8) | 134, 100.0 (96.6 – 100.0) |
|  | DENV infection (German travelers), panel A | 12 | 12, 100.0 (71.8 – 100.0) | 12, 100.0 (71.8 – 100.0) |
|  | DENV infection (Lao PDR residents), S1 | 35 | 31, 88.6 (73.4 – 96.1) | 35, 100.0 (88.2 – 100.0) |
|  | DENV infection (Lao PDR residents), S2 | 35 | 16, 45.7 (30.5 – 61.8) | 35, 100.0 (88.2 – 100.0) |
|  | DENV infection (Colombia residents), S1 | 12 | 10, 83.3 (54.0 – 96.5) | 12, 100.0 (71.8 – 100.0) |
|  | DENV infection (Colombia residents), S2 | 12 | 5, 41.7 (19.3 – 68.1) | 12, 100.0 (71.8 – 100.0) |
|  | WNV infection (Canada residents) | 17 | 17, 100.0 (78.4 – 100.0) | 17, 100.0 (78.4 – 100.0) |
|  | YF vaccinees (German travelers) | 91 | 91, 100.0 (95.1 – 100.0) | 91, 100.0 (95.1 – 100.0) |
|  | TBEV NS1 IgG positive | 2 | 2, 100.0 (29.0 – 100.0) | 2, 100.0 (29.0 – 100.0) |
|  | CHIKV infection (WHO IgG standard, 1502/19) | 1 | 1, 100.0 (16.7 – 100.0) | 1, 100.0 (16.7 – 100.0) |
|  | *Plasmodium* infection (residual samples) | 54 | 53, 98.1 (89.3 – 100.0) | 54, 100.0 (92.1 – 100.0) |
|  | Complete panel | 667 | 604, 90.6 (88.1 – 92.6) | 667, 100.0 (99.3 – 100.0) |

**Table 2C.** WNV lin1 (NS1) IgG FcyR ELISA validation data. ^1^2/3 samples testing negative in ELISA are WNV PCR positive, ^2^for the 4 samples testing positive in ELISA, presence of anti-flavivirus IgG was confirmed by independent serological testing (**Supplemental** Table 2). Abbreviations: w/o: without, w/: with, DENV: Dengue virus, ZIKV: Zika virus, WNV: West Nile virus, YF: Yellow fever, TBEV: Tick-borne encephalitis virus, CHIKV: Chikungunya virus, CV: coefficient of variation, CI: confidence interval, WHO: World Health Organization, N: number of sera or patients tested.

| **WNV lin1 (NS1) IgG FcγR ELISA** |  |  | **w/o *SE*** | **w/ *SE*** |
|---|---|---|---|---|
| **Assay reagents** | HRP-labeled antigen |  | WNV lineage 1 NS1 | WNV lineage 1 NS1 |
|  | Specificity Enhancer (*SE*) |  | none | DENV1-4/ZIKV/JEV/USUV/YFV NS1 |
| **Reproducibility** | Intra-assay CV |  | ≤ 5% | ≤ 5% |
|  | Inter-assay CV |  | ≤ 10% | ≤ 10% |
| **Sensitivity** | **Serum panel** | **N** | **n positive, % sensitivity (95% CI)** | **n positive, % sensitivity (95% CI)** |
|  | WNV infection (Canada residents) | 24 | 24, 100.0 (83.7 – 100.0) | 24, 100.0 (83.7 – 100.0) |
|  | WNV infection (Greece residents) | 18 | 17, 94.4 (72.3 – 100.0) | 15, 83.3 (60.0 – 95.0)<sup>1</sup> |
| **Specificity** | **Serum panel** | **N** | **n negative, % specificity (95% CI)** | **n negative, % specificity (95% CI)** |
|  | Healthy donors (Germany residents) | 35 | 35, 100.0 (88.2 – 100.0) | 35, 100.0 (88.2 – 100.0) |
|  | Healthy donors (Ukraine residents) | 200 | 200, 100.0 (97.7 – 100.0) | 200, 100.0 (97.7 – 100.0) |
|  | Healthy donors (Lao PDR residents) | 5 | 4, 80.0 (36.0 – 98.0) | 5, 100.0 (51.1 – 100.0) |
|  | Healthy donors (Colombia residents) | 9 | 5, 55.6 (26.6 – 81.2) | 9, 100.0 (65.5 – 100.0) |
|  | Healthy donors (Ghana residents) | 35 | 33, 94.3 (80.4 – 99.4) | 34, 97.1 (84.2 – 100.0) |
|  | DENV infection (German travelers, residual samples) | 8 | 8, 100.0 (62.8 – 100.0) | 8, 100.0 (62.8 – 100.0) |
|  | DENV infection (Lao PDR residents), S1 | 35 | 33, 94.3 (80.4 – 99.4) | 35, 100.0 (88.2 – 100.0) |
|  | DENV infection (Lao PDR residents), S2 | 35 | 17, 48.6 (33.0 – 64.4) | 35, 100.0 (88.2 – 100.0) |
|  | DENV infection (Colombia residents), S1 | 3 | 2, 66.7 (20.2 – 94.4) | 3, 100.0 (38.2 – 100.0) |
|  | DENV infection (Colombia residents), S2 | 8 | 1, 12.5 (0.1 – 49.2) | 8, 100.0 (62.8 – 100.0) |
|  | ZIKV infection (German travelers) | 7 | 7, 100.0 (59.6 – 100.0) | 7, 100.0 (59.6 – 100.0) |
|  | YF vaccinees (German travelers) | 91 | 90, 98.9 (93.4 – 100.0) | 91, 100.0 (95.1 – 100.0) |
|  | TBEV NS1 IgG positive | 2 | 2, 100.0 (29.0 – 100.0) | 2, 100.0 (29.0 – 100.0) |
|  | CHIKV infection (WHO IgG standard, 1502/19) | 1 | 1, 100.0 (16.7 – 100.0) | 1, 100.0 (16.7 – 100.0) |
|  | *Plasmodium* infection (residual samples) | 54 | 47, 87.0 (75.3 – 93.9) | 50, 92.6 (81.9 – 97.6)<sup>2</sup> |
|  | Complete panel | 528 | 485, 91.9 (89.2 – 93.9) | 523, 99.1 (97.7 – 99.7) |

**Table 2D.** WNV lin2 (NS1) IgG FcyR ELISA validation data. ^1^IgG seroconversion detected in 8/8 patients, ^2^the 2 samples testing negative in ELISA are WNV PCR positive, ^3^for the 2 samples testing positive in ELISA, presence of anti-flavivirus IgG was confirmed by independent serological testing (**Supplemental** Table 2). Abbreviations: w/o: without, w/: with, DENV: Dengue virus, ZIKV: Zika virus, WNV: West Nile virus, YF: Yellow fever, TBEV: Tick-borne encephalitis virus, CHIKV: Chikungunya virus, CV: coefficient of variation, CI: confidence interval, WHO: World Health Organization, N: number of sera or patients tested.

| **WNV lin2 (NS1) IgG FcγR ELISA** |  |  | **w/o *SE*** | **w/ *SE*** |
|---|---|---|---|---|
| **Assay reagents** | HRP-labeled antigen |  | WNV lineage 2 NS1 | WNV lineage 2 NS1 |
|  | Specificity Enhancer (*SE*) |  | none | DENV1-4/ZIKV/JEV/USUV/YFV NS1 |
| **Reproducibility** | Intra-assay CV |  | ≤ 5% | ≤ 5% |
|  | Inter-assay CV |  | ≤ 10% | ≤ 10% |
| **Sensitivity** | **Serum panel** | **N** | **n positive, % sensitivity (95% CI)** | **n positive, % sensitivity (95% CI)** |
|  | WNV infection (Ukraine residents) | 8 | 8, 100.0 (62.8 – 100.0) | 8, 100.0 (62.8 – 100.0)<sup>1</sup> |
|  | WNV infection (Canada residents) | 24 | 24, 100.0 (83.7 – 100.0) | 24, 100.0 (83.7 – 100.0) |
|  | WNV infection (Greece residents) | 18 | 17, 94.4 (72.3 - 100.0) | 16, 88.9 (65.9 – 98.1)<sup>2</sup> |
|  | WNV infection (Slovenia residents) | 3 | 3, 100.0 (38.2 – 100.0) | 3, 100.0 (38.2 – 100.0) |
| **Specificity** | **Serum panel** | **N** | **n negative, % specificity (95% CI)** | **n negative, % specificity (95% CI)** |
|  | Healthy donors (Germany residents) | 35 | 35, 100.0 (88.2 – 100.0) | 35, 100.0 (88.2 – 100.0) |
|  | Healthy donors (Ukraine residents) | 200 | 200, 100.0 (97.7 – 100.0) | 200, 100.0 (97.7 – 100.0) |
|  | Healthy donors (Lao PDR residents) | 5 | 5, 100.0 (51.1 – 100.0) | 5, 100.0 (51.1 – 100.0) |
|  | Healthy donors (Colombia residents) | 9 | 5, 55.6 (26.6 – 81.2) | 9, 100.0 (65.5 – 100.0) |
|  | Healthy donors (Ghana residents) | 35 | 34, 97.1 (84.2 – 100.0) | 35, 100.0 (88.2 – 100.0) |
|  | DENV infection (German travelers, residual samples) | 8 | 8, 100.0 (62.8 – 100.0) | 8, 100.0 (62.8 – 100.0) |
|  | DENV infection (Lao PDR residents), S1 | 35 | 34, 97.1 (84.2 – 100.0) | 35, 100.0 (88.2 – 100.0) |
|  | DENV infection (Lao PDR residents), S2 | 35 | 18, 51.4 (35.6 – 67.0) | 35, 100.0 (88.2 – 100.0) |
|  | DENV infection (Colombia residents), S1 | 3 | 3, 100.0 (38.2 – 100.0) | 3, 100.0 (38.2 – 100.0) |
|  | DENV infection (Colombia residents), S2 | 8 | 2, 25.0 (6.3 – 59.9) | 8, 100.0 (62.8 – 100.0) |
|  | ZIKV infection (German travelers) | 7 | 7, 100.0 (59.6 – 100.0) | 7, 100.0 (59.6 – 100.0) |
|  | YF vaccinees (German travelers) | 91 | 90, 98.9 (93.4 – 100.0) | 91, 100.0 (95.1 – 100.0) |
|  | TBEV NS1 IgG positive | 2 | 2, 100.0 (29.0 – 100.0) | 2, 100.0 (29.0 – 100.0) |
|  | TBEV infection (Slovenia residents) | 15 | 15, 100.0 (76.1 – 100.0) | 15, 100.0 (76.1 – 100.0) |
|  | CHIKV infection (WHO IgG standard, 1502/19) | 1 | 1, 100.0 (16.7 – 100.0) | 1, 100.0 (16.7 – 100.0) |
|  | *Plasmodium* infection (residual samples) | 54 | 49, 90.7 (79.7 – 96.4) | 52, 96.3 (86.7 – 99.7)<sup>3</sup> |
|  | Complete panel | 543 | 508, 93.6 (91.1 – 95.4) | 541, 99.6 (98.6 – 100.0) |

### Specificity (flavivirus subgroup): cross-reactivity and suppression by SE

35 serum samples collected between 14 and 42 days post onset of symptoms from PCR-confirmed DF patients from Lao PDR, a DENV endemic country with no reported circulation of either ZIKV or WNV at the time of sampling, showed remarkably strong cross-reactivity (≥ 49%) in the ZIKV and WNV (NS1) IgG FcγR ELISAs w/o *SE* (**Figure 1**, **Table 2B-D** (panel “DENV infection (Lao PDR residents), S2”)). Cross-reactivity was mainly observed in samples highly reactive in the DENV (NS1) IgG FcγR ELISA w/o *SE* (**Supplemental Figure 2**) and could be completely eliminated by the addition of *SE* (**Table 2B-D, Figure 1**). In contrast, cross-reactivity of serum samples from European individuals having received a heterologous flavivirus vaccine or contracted a flavivirus infection abroad was absent or low even w/o *SE* (**Table 2A-D**). For example, 24/24 baseline samples from DENV-naïve European individuals intending to receive DENV vaccination tested negative in the DENV (NS1) IgG FcγR ELISA w/o *SE* (**Figure 2B**, **Table 2A** (panel “DENV naïve vaccinees (Germany residents), S0 (baseline)”), although 21/24 individuals reported previous flavivirus vaccination(s) and serological evidence of previous flavivirus contact could be detected in 15 of those 21 individuals by ELISA and IIFT (data not shown). Importantly, no cross-reactivity of serum samples from the acute, early and late convalescent phases of 15 TBE patients (**Supplemental Table 1**) was observed in the WNV lin2 (NS1) IgG FcγR ELISA w/o *SE* (**Table 2D**).

**Figure 1.**
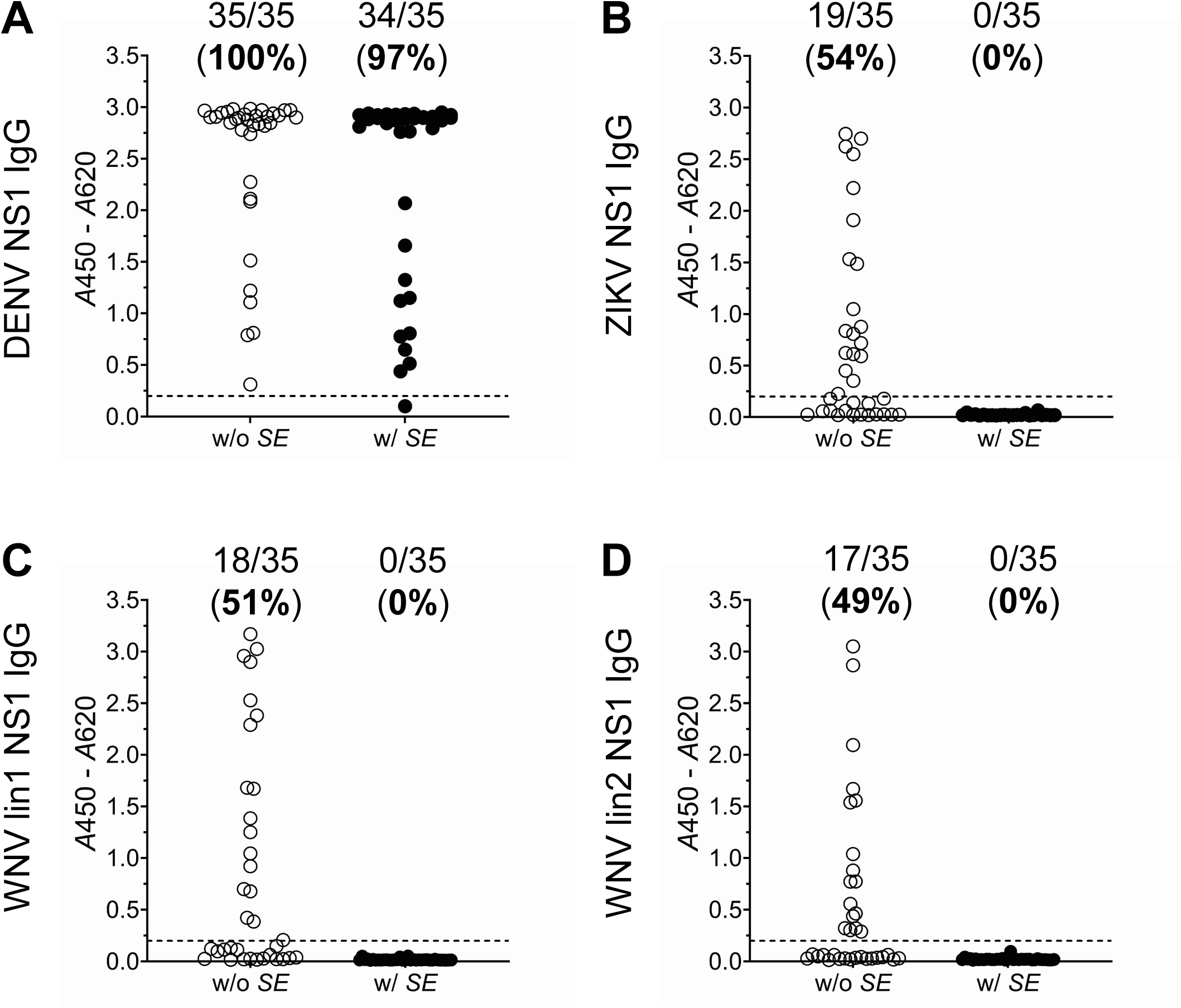
**Reactivity of DF patient sera in flavivirus IgG Fc**y**R ELISAs and elimination of cross-reactivity with *SE*.** Serum samples from 35 Laotian patients with a PCR-confirmed DENV infection, collected between day 14 and 42 post onset of symptoms, were analyzed with the **(A)** DENV, **(B)** ZIKV, **(C)** WNV lin1, and **(D)** WNV lin2 (NS1) IgG FcyR ELISAs in absence (w/o) and presence (w/) of the assay-specific *SE*. *A*: absorbance, dotted line indicates assay cut-off, n/N positive (percentage).

**Figure 2.**
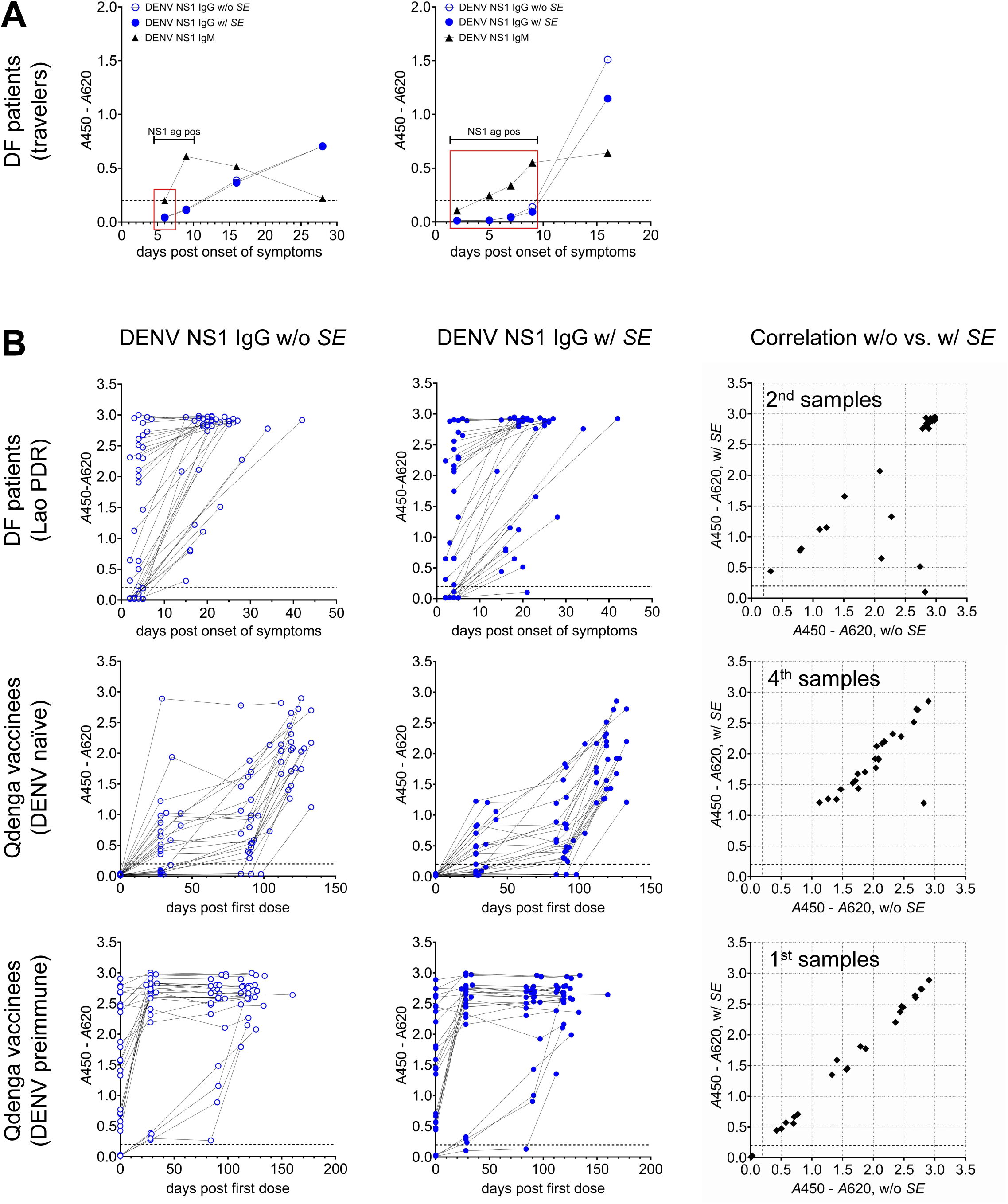
**DENV (NS1) IgG Fc**y**R ELISA results w/o and w/ *SE*** for longitudinal serum samples from **(A)** DF patients (German travelers, red box indicates DENV PCR-positive samples, capped horizontal line indicates period of NS1 antigen positivity) and **(B)** DF patients (Lao PDR, N=35), DENV-naïve Qdenga vaccinees (Germany, N=24), and DENV preimmune Qdenga vaccinees (Germany, N=25). *A*: absorbance, ag: antigen, pos: positive, dotted line indicates assay cut-off.

### Specificity (non-flaviviral infections subgroup)

Specificity of flavivirus IgG ELISAs has been found to be challenged not only by flavivirus cross-reactivity, but also by previous contact with other, phylogenetically unrelated, pathogens like SARS-CoV-2 (18, 29–31) and Plasmodium *spp*. (32). Notably, 19/19 serum samples from German donors with a high anti-SARS-CoV-2 IgG titer (**Supplemental Table 1**) tested negative in the DENV (NS1) IgG FcγR ELISA w/o *SE* (**Table 2A**). A possible cause of malaria-associated interference in serological assays is reactivity of antibodies elicited against histidine-rich epitopes with His-tagged recombinant antigens (33). Nevertheless, 53/54 and 54/54 *Plasmodium* IgG IIFT positive serum samples (**Supplemental Table 1)** tested negative in the ZIKV (NS1) IgG FcγR ELISA w/o and w/ *SE*, respectively (**Table 2B, Supplemental Table 2),** although the assay is employing recombinant, His-tagged ZIKV NS1 as antigen that is recognized by a commercially available anti-His monoclonal antibody (data not shown). Given the widely overlapping geographic distribution of *Plasmodium,* DENV, and WNV, previous contact of the donors with those viruses cannot be ruled out. Indeed, 6/54 samples reacted positive in the DENV (NS1) IgG FcγR ELISA w/ *SE* (but not in the WNVlin1/2 (NS1) IgG FcγR ELISAs w/ *SE*), 3/54 reacted positive in the WNV lin1 (NS1) IgG FcγR ELISA w/ *SE* (but not in the DENV (NS1) IgG FcγR ELISA w/ *SE*), and 1/54 samples reacted positive in both assays (**Tables 2A, C, D, Supplemental Table 2**). In all samples testing positive in the DENV (NS1) and/or the WNVlin1/2 (NS1) IgG FcγR ELISAs, anti-flavivirus IgG antibodies were also detectable by alternative binding antibody assays based on different test principles (IIFT, indirect ELISA) and/or employing different antigens (native virus, envelope protein) (**Supplemental Table 2**).

### Sensitivity (DENV (NS1) IgG FcγR ELISA w/o and w/ SE)

The assays reliably detected anti-DENV NS1 IgG in convalescent-phase samples from PCR-confirmed DF patients, including both travelers and residents of endemic countries (**Table 2A, Figure 2A**, **Figure 2B**). They also detected seroconversion in 100% (24/24) of samples from previously DENV-naïve individuals who had completed a DENV vaccination regimen (**Table 2A, Figure 2B**). Furthermore, anti-DENV NS1 IgG antibodies were detected in 84% (21/25) of baseline samples of self-reported DENV preimmune individuals intending to receive a DENV vaccine (**Table 2A, Figure 2B**) and in 68% (32/47) of PCR-positive early acute-phase samples from individuals from DENV endemic countries with high rates of secondary infections (**Table 2A**, for Lao PDR: **Figure 2B**). Importantly, addition of *SE* did not result in a significant loss of assay sensitivity (**Table 2A, Figure 2**): no or only a minor effect on obtained *A*450-*A*620 values was observed in the vast majority of sera from German residents having contracted DF abroad or having received a DENV vaccination (**Figure 2A, 2B)**, even if they reported previous JEV, TBEV, or YFV vaccinations. However, DENV (NS1) IgG FcγR signal heights were noticeably (≥ 42 %) decreased upon addition of *SE* in 4/35 samples obtained from Laotian PCR-confirmed DF patients between day 14 and 42 post onset of symptoms (**Figure 2B**). Functionality testing of the individual components of the *SE* on these 4 samples revealed that suppression was mainly mediated by the NS1 of JEV (**Supplemental Figure 3A**), the only flavivirus besides DENV reportedly circulating at significant levels in Lao PDR at the time of sampling (34, 35). Indeed, all four samples showed high reactivity with HRP-labeled JEV NS1 that could not be suppressed below cut-off by the addition of an excess of unlabeled DENV/ZIKV/WNV/USUV/YFV NS1 proteins (**Supplemental Figure 3B**).

### Sensitivity (ZIKV (NS1) IgG FcγR ELISA w/o and w/ SE)

Anti-ZIKV NS1 IgG seroconversion was reliably observed in convalescent phase serum samples from returning European travelers with a PCR-confirmed ZIKV infection (**Table 2B, Figure 3A**), and ZIKV NS1 IgG antibodies were detected in ZIKV PRNT positive serum samples from adults (8/9 positive w/o *SE*, 7/9 positive w/ *SE*) and children (5/5 positive w/o and w/ *SE*) from a DENV endemic country (Brazil) (**Table 2B, Figure 3B**). A considerable drop of signal heights in the ZIKV (NS1) IgG FcγR ELISA was induced by the *SE* in ZIKV PRNT positive sera from adult Brazilian donors that tested strongly positive in the DENV (NS1) IgG FcγR ELISA w/o and w/ *SE*, but not in sera from Brazilian children with no apparent previous contact to DENV (**Figure 3B**).

**Figure 3.**
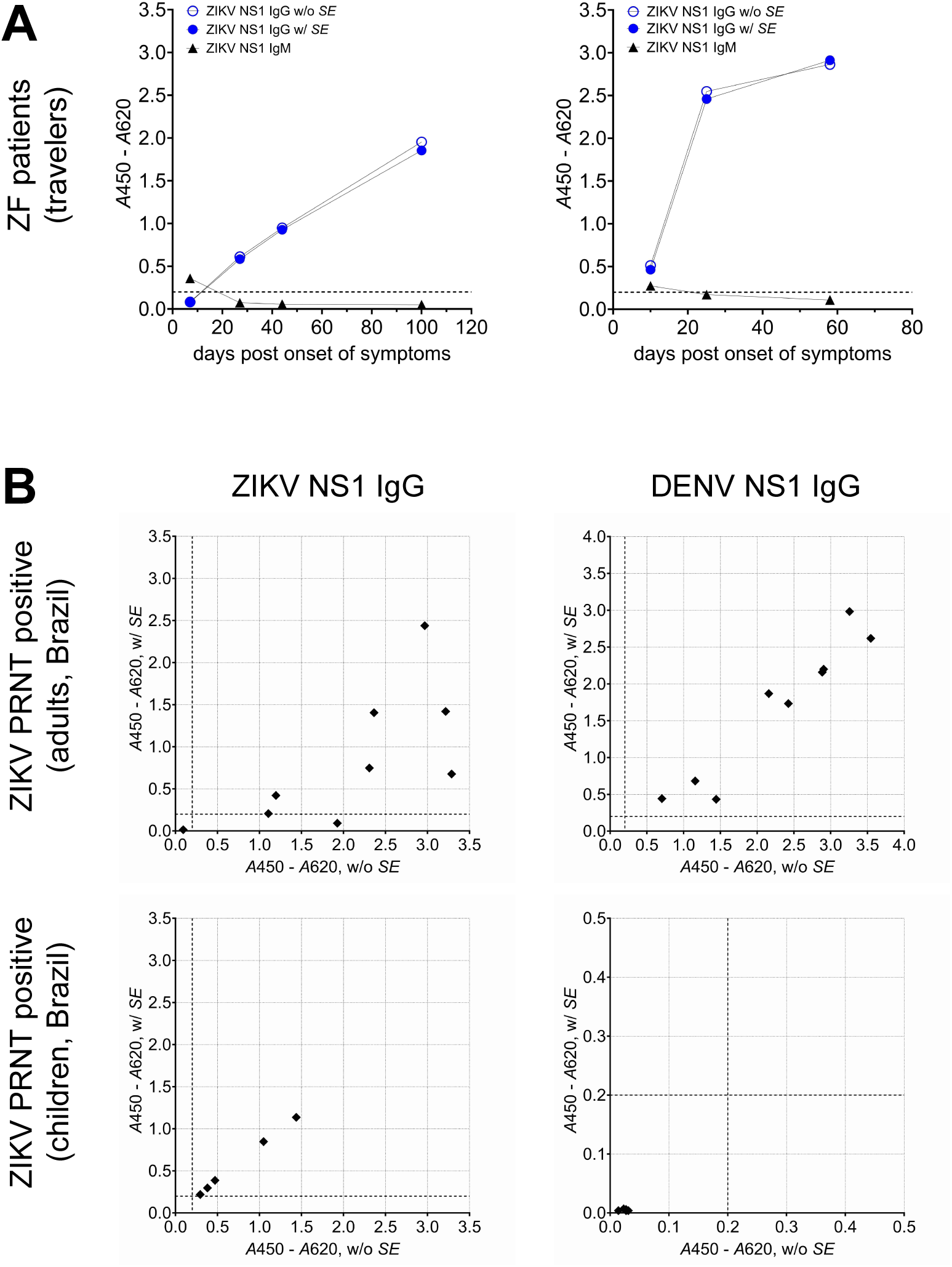
**ZIKV (NS1) IgG Fc**y**R ELISA results w/o and w/ *SE*** for **(A)** longitudinal serum samples from ZF patients (German travelers) and **(B)** ZIKV PRNT positive serum samples from Brazil; samples were analyzed with the ZIKV and DENV (NS1) IgG FcyR ELISAs w/o and w/ *SE*. *A*: absorbance, dotted line indicates assay cut-off.

### Sensitivity (WNVlin1/2 (NS1) IgG FcγR ELISA w/o and w/ SE)

Anti-WNV NS1 IgG antibodies were reliably detected in early-convalescent-and convalescent-phase sera from WNF patients from European countries (Ukraine, Slovenia, Greece) as well as from Canada (**Table 2C, Table 2D, Figure 4A-D**). No significant influence of the *SE* on signal intensities was observed in the majority of WNF patient sera (**Figure 4A**, **Figure 4B**, **Figure 4D**).

**Figure 4.**
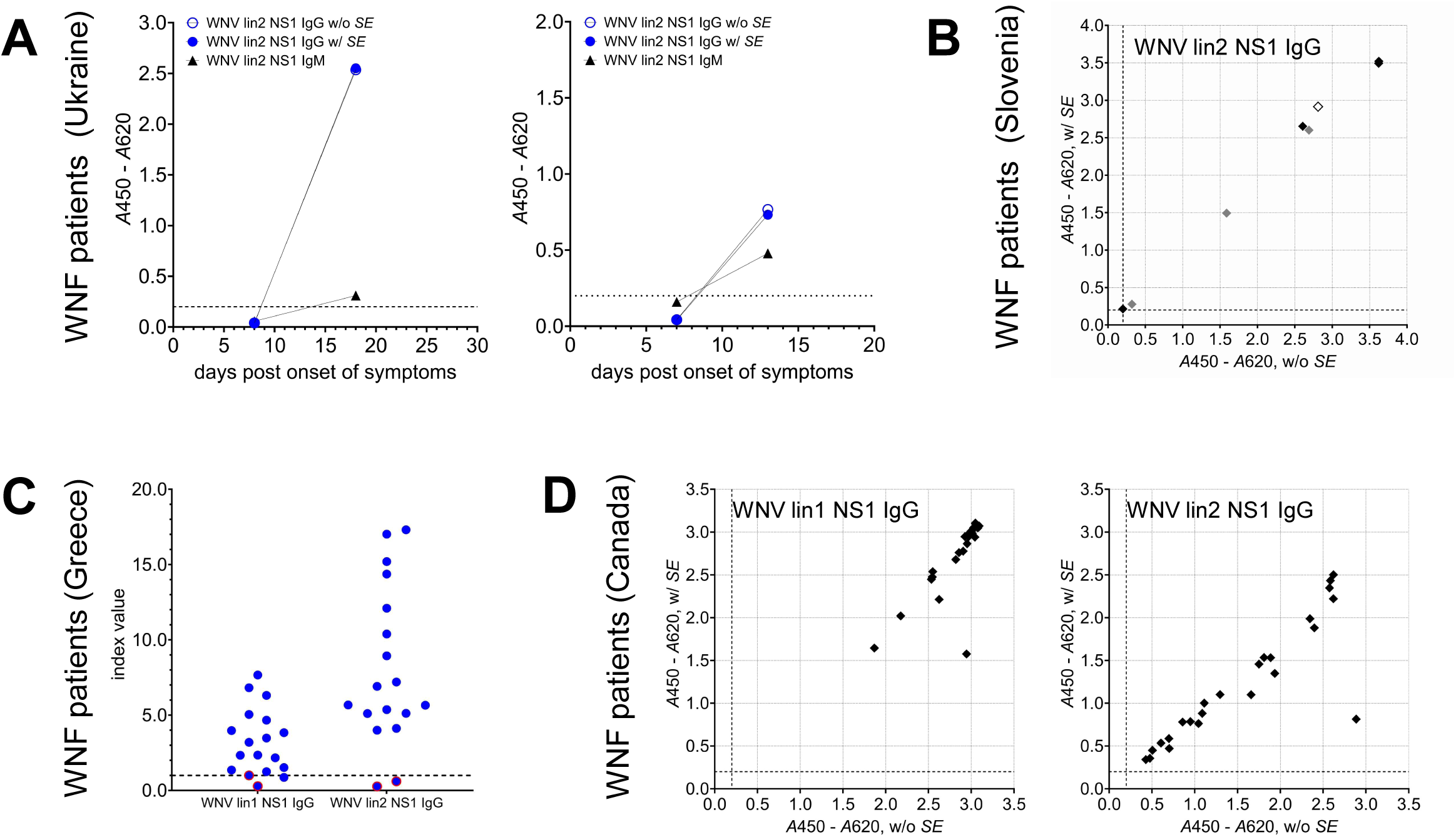
**WNV (NS1) IgG Fc**y**R ELISA results** for **(A)** longitudinal serum samples from Ukrainian WNF patients, **(B)** longitudinal samples from 3 Slovenian patients with PCR-confirmed WNV infection (samples with matching colors were taken from the same patient at different timepoints), **(C)** samples from 18 Greek WNF patients, antigens: WNV lin1 NS1 and WNV lin2 NS1, tested w/ *SE*, WNV PCR-positive samples are marked in red, **(D)** WNV PRNT positive samples from Canada (N=24), antigens: WNV lin1 NS1 and WNV lin2 NS1; *A*: absorbance, index value: absorbance values normalized to cut-off, dotted line indicates assay cut-off.

### The discriminatory power of Flavivirus (NS1) IgG FcγR SE ELISAs: proof-of-concept serological differentiation of WNV lineages

Considering the spatiotemporal distribution of WNV lineages, European patients most likely had been exposed to WNV lineage 2, while infections of Canadian patients were caused by WNV lineage 1 (36). While in general inter-lineage cross-detection was observed when working with either WNV lineage 1 or 2 NS1 as ELISA antigen (**Table 2C, Table 2D**), significant differences (paired t test, p < 0.0001) in signal heights were observed when comparing homologous to heterologous detection (**Figure 4C**, **Figure 4D**). Therefore, we assessed if serological discrimination of previous contact with WNV lineage 1 and 2, respectively, can be achieved. Indeed, by combination of WNV lin1 NS1 HRP-labeled antigen and WNV lin2 NS1 as competitor and vice versa, lineage-specific detection could be established in a small exemplary sample set comprising WNV IgG positive serum samples from Canada and Europe, respectively (**Figure 5B**).

**Figure 5.**
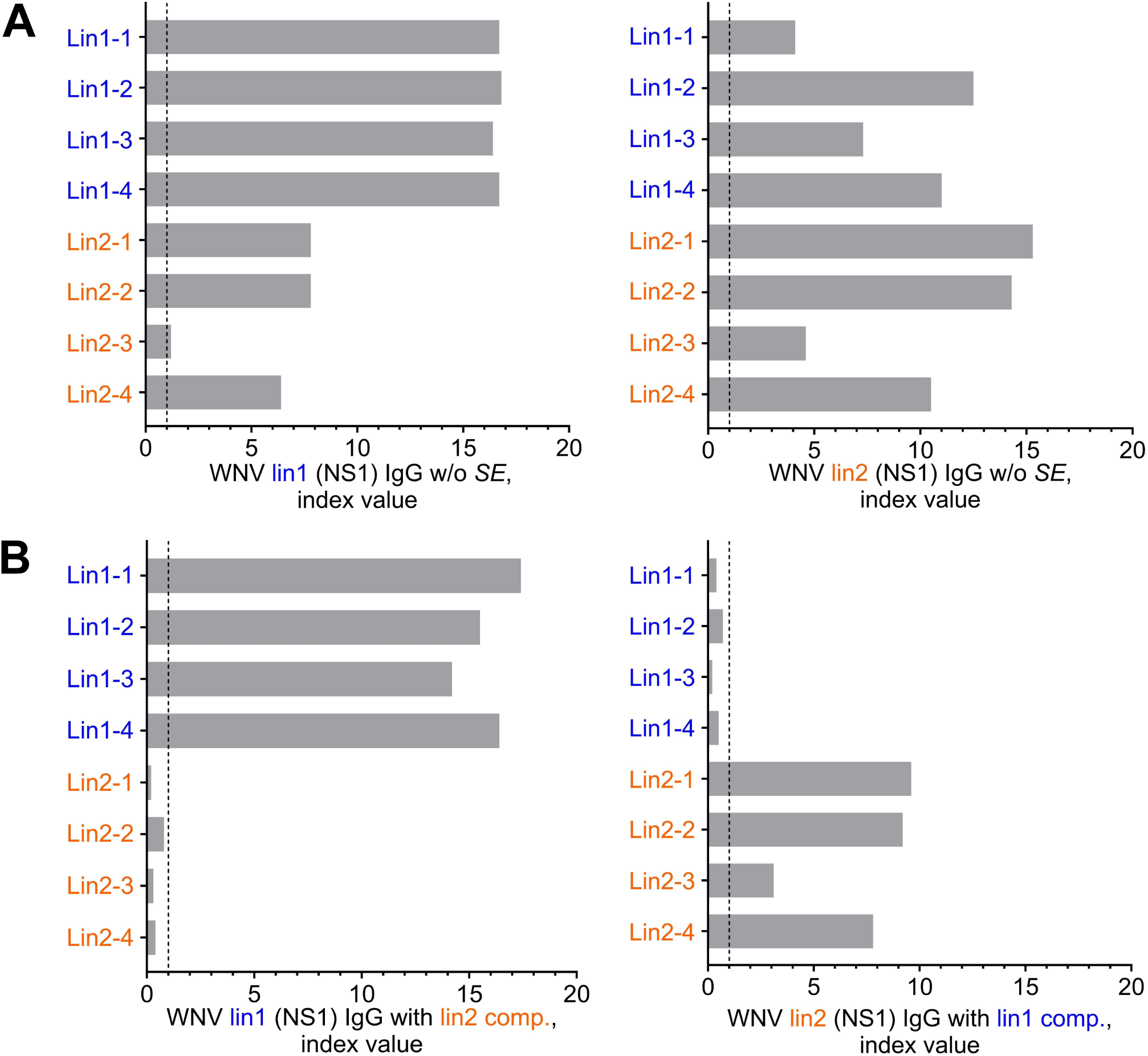
Proof-of-concept: differential detection of IgG antibodies elicited by exposure to WNV lineages 1 and 2. **(A)** Reactivity of WNV IgG positive serum samples from Canada (lineage 1) and Europe (presumable lineage 2), respectively, in WNV lin1/2 (NS1) IgG FcyR ELISA w/o *SE*, **(B)** suppression of lineage cross-detection by addition of recombinant heterologous competitor molecule; index value: absorbance values normalized to cut-off, dotted line indicates assay cut-off.

## DISCUSSION

### Flavivirus NS1 as ELISA antigen

Previously, we established HRP-labeled ZIKV NS1 as antigen in a prototypic ZIKV (NS1) IgG FcγR ELISA (20). When validated using longitudinal serum samples from PCR-confirmed Zika fever patients from Brazil and *a priori* ZIKV IgG negative serum samples from DENV endemic countries, this assay was found to be as sensitive, but significantly more specific than a commercial, CE-marked NS1-based indirect ELISA (20). The observed superior specificity is most likely caused by two factors: 1) the use of soluble NS1 protein ensuring preservation of conformational epitopes and avoidance of artificial exposure of hydrophobic regions and 2) the *Hs*FcγR-based capture technology which may further reduce the influence of low-avidity antigen/antibody interactions. However, even under these conditions, sera from DENV-endemic regions remained a major source of cross-reactivity (20), highlighting the need for further refinement of NS1-based serological approaches.

### Flavivirus (NS1) IgG FcγR SE ELISAs

In this study, we developed NS1-based IgG FcγR ELISAs for three medically relevant mosquito-borne flaviviruses: DENV, ZIKV, and WNV. To reduce serological cross-reactivity, we incorporated a target-specific Specificity Enhancer (*SE*) reagent consisting of recombinant, unlabeled homologues derived from other flaviviruses with known cross-reactive potential, a technological concept that has been successfully applied previously in EDIII-based flavivirus IgG ELISAs (20, 24). Validation of the assays demonstrated very good reproducibility as well as high clinical sensitivity and specificity; cross-reactive responses caused by antibodies binding to conserved NS1 epitopes from non-target flaviviruses were efficiently suppressed. The influence of *SE* on signals from true-positive samples varied considerably depending on infection or vaccination history, underlining the complexity of flavivirus immune imprinting and its impact on binding antibody detection. Notably, the system enabled discrimination even between IgG antibodies elicited by contact with very closely related viruses, such as different WNV lineages, despite the extraordinarly high sequence identity at the amino acid level. This observation is in line with previous findings demonstrating differential antibody reactivities to NS1 proteins of tick-borne encephalitis virus (TBEV) strains (37).

The assays and reagents established in this study constitute a modular toolkit for the serological characterization of previous flavivirus exposure. By systematically comparing reactivities in absence and presence of *SE* and analyzing the contribution of individual *SE* components to signal modulation, it is possible to derive a differentiated, plausible profile of prior flavivirus contacts. This strategy was exemplified using the Lao serum panel, in which distinct patterns of reactivity allowed reconstruction of probable exposure histories.

### Limitations of the study/the developed assays

Several limitations of this study and of the developed assays should be considered: First of all, while plaque reduction neutralization testing (PRNT) is widely accepted as the gold standard for the detection of flavivirus-neutralizing antibodies, at least in primary infections (38), no comparable reference method exists for the highly specific detection of flavivirus-binding IgG antibodies. Consequently, the analytical sensitivity of highly specific NS1-based assays may appear reduced when compared with serological tests picking up cross-reactive responses that are often boosted in acute infections (3). In general, studies on flavivirus antibody binding assays are hampered by incomplete information on extent and timeline of previous flavivirus exposures. To mitigate this challenge, samples generating debatable results in this study were, if possible, subjected to additional serological testing providing complementary information like indirect immunofluorescence analysis (IIFT) or ELISAs based on alternative antigens like EDIII. Furthermore, anti-NS1 IgG seroconversion has been observed to occur slightly later than anti-EP IgG seroconversion (13, 39) which might become relevant when analyzing acute phase serum samples. Therefore, parallel testing of acute phase samples for target-specific IgM antibodies is highly recommended (**Figure 2A**, **Figure 3A)**. Applicability of NS1-based flavivirus IgG ELISAs for the evaluation of vaccine responses strongly depends on the target pathogen: while no or only low levels of anti-NS1 IgG antibodies are induced upon vaccination with inactivated TBEV vaccines (40–42), robust induction of anti-NS1 IgG antibodies is observed upon vaccination with the live-attenuated DENV vaccine Qdenga (TAK-003) ((43) and this study). Preliminary data using a prototypic YFV (NS1) IgG FcγR ELISA revealed anti-NS1 IgG seroconversion in several, but not all, tested individuals having received an attenuated YFV vaccine (data not shown). This observation is in line with a recent study demonstrating absent or low level RNAemia, but nevertheless induction of anti-NS1 antibodies indicative of viral replication, in YF-17D vaccinees (44).

Due to the limited availability of comprehensive, well-characterized serum panels from individuals with PCR-confirmed JEV and USUV infections, the respective extent of cross-reactivity and its suppression by the *SEs* could not be systematically assessed in the present study but will be addressed in future investigations.

Specific caveats result from the employed assay technology: although the *Hs*FcγRIIA H131 variant used as capture molecule binds to all four IgG subclasses, its affinity is highest for IgG1 and IgG3 (the predominant subclasses induced by natural DENV infection (45, 46)), but significantly lower for IgG2 and IgG4 (47). Furthermore, masking of relevant epitopes might occur due to direct covalent coupling of the recombinant antigen to HRP. As exemplified above, the effect of individual *SE* components can be highly selective (**Supplemental Figure 3**). Therefore, antibodies induced by unrepresented flaviviruses might not be efficiently outcompeted by the current *SE* formulations. However, the modular design of the technology platform allows straightforward adaptation to emerging diagnostic needs through inclusion of additional NS1 competitor molecules.

### Conclusion and perspectives

In conclusion, the NS1-based IgG FcγR/*SE* ELISA platform represents a valuable tool for high-resolution analysis of flavivirus IgG seroprevalence, particularly in sera originating from complex endemic settings with overlapping virus circulation, as exemplified in recent field applications (48). The combination of FcγR-mediated antibody capture with targeted suppression of cross-reactive signals enables improved differentiation of flavivirus-specific immune responses beyond the capacity of conventional serological assays.

The modular “toolbox” design allows straightforward adaptation to additional medically relevant flaviviruses such as YFV, JEV, TBEV, Saint Louis Encephalitis virus (SLEV), or Alkhurma virus (ALKV), and the underlying principle is transferable to other antigenically related virus groups, for example alphaviruses (e.g. CHIKV, O’nyong-nyong virus (ONNV), Mayaro virus (MAYV)). Moreover, human Fcγreceptors promiscuously bind to IgG from multiple mammalian species, including rodents (49), ruminants (26), equids, camelids, and non-human primates (data not shown), opening up new possibilities for seroepidemiological studies in a One Health framework.

## Data Availability

All data produced in the present study are available upon reasonable request to the authors.

## ACKNOWLEDGMENTS

We thank M. Loeb and P. Singh for providing serum samples from Canada, L. J. Maya Amaya and G. E. Rivera Salcedo for providing serum samples from Colombia, and M. Mackroth for providing serum samples from YFV vaccinees. Furthermore, we thank Y. Winkelmann and J. Hansen for expert technical assistance.

## List of abbreviations (in alphabetical order)

A: absorbance
Ag: antigen
ALKV: Alkhurma virus
AUC: area under curve
BNITM: Bernhard Nocht Institute for Tropical Medicine
BSA: bovine serum albumin
cDNA: complementary DNA
CE: Conformité Européenne
CHIKV: Chikungunya virus
CI: Confidence interval
CV: Coefficient of variation
DENV: Dengue virus
DF: Dengue fever
Dpo: days post onset of symptoms
EDIII: Envelope protein domain III
ELISA: Enzyme-linked Immunosorbent Assay
EP: Envelope protein
ERDF: European Regional Development Fund
FcγR: Fcy receptor
FcµR: Fcµ receptor
HRP: Horseradish peroxidase
IgG: immunoglobulin G
IgM: immunoglobulin M
IIFT: indirect immunofluorescence testing
JEV: Japanese encephalitis virus
Lao PDR: Lao People’s Democratic Republic
Lin: lineage
MAYV: Mayaro virus
NIBSC: National Institute for Biological Standards and Control
NTA: nitrilotriacetic acid
NS1: nonstructural protein 1
ONNV: O’nyong-nyong virus
PCR: Polymerase Chain Reaction
PEI: Paul Ehrlich Institute
PRNT: Plaque reduction neutralization testing
RKI: Robert Koch Institute
RNA: ribonucleic acid
ROC: Receiver operating characteristic
SARS-CoV-2: Severe Acute Respiratory Syndrome Corona virus 2
SLEV: Saint Louis Encephalitis virus
SE: Specificity Enhancer
SEC: size-exclusion chromatography
TBEV: Tick-borne encephalitis virus
TMB: tetramethylbenzidine
USUV: Usutu virus
w/: with
w/o: without
WHO: World Health Organization
WNF: West Nile fever
WNV: West Nile virus
YFV: Yellow fever virus
ZIKV: Zika virus
ZF: Zika fever

## SUPPLEMENTAL TABLE CAPTIONS

**Supplemental Table 1: Human serum samples.** N: number of patients/donors, n: number of samples. The table summarizes information of complete sample panels; subsets used for individual assay validations are stated in tables 2A-D; NIBSC: National Institute for Biological Standards and Control, BNITM: Bernhard Nocht Institute for Tropical Medicine, PEI: Paul Ehrlich Institute, RKI: Robert Koch Institute, dpo: days post onset of symptoms

**Supplemental Table 2. Analysis of *Plasmodium* IgG IIFT positive serum samples.** Samples (n=54) were tested with the DENV, ZIKV, and WNV (NS1) IgG FcγR ELISAs in absence (w/o) and presence (w/) of the assay-specific *SE* (cut-off *A*450 – *A*620 = 0.200, green: negative, orange: positive). Positive samples were subjected to confirmatory testing with flavivirus IgG IIFT and/or ELISA (Panbio DENV IgG ELISA, Focus WNV IgG ELISA, WNV Envelope Protein (EP) IgG FcγR ELISA (employing recombinant antigen comprising EP domains DI, DII, and DIII)).

## SUPPLEMENTAL FIGURE CAPTIONS

**Supplemental Figure 1. Detection range and ROC analyses (example DENV (NS1) IgG Fc**γ**R ELISA). (A)** Detection range. Serial dilutions of anti-DENV1-4 NS1 rabbit hyperimmune serum were analyzed with the DENV (NS1) IgG FcγR ELISA; dotted line indicates assay cut-off (*A*450 – *A*620 = 0.2). **(B) – (E)** ROC analysis was performed for paired serum samples from 35 Laotian patients with a PCR-confirmed DENV infection collected either between day 2 and 7 (median day 4) and between day 14 and 42 (median day 20) post onset of symptoms, respectively, and 42 serum samples from healthy European donors. *A*: absorbance, AUC: area under curve, CI: confidence interval.

Supplemental Figure 2. Correlation of DENV (NS1) IgG FcγR ELISA w/o *SE* results and (A) ZIKV, (B) WNV lin1, (C) WNV lin2 (NS1) IgG FcγR ELISA w/o *SE* results for serum samples from 35 Laotian patients with a PCR-confirmed DENV infection, collected between day 14 and 42 post onset of symptoms. *A*: absorbance, dotted line indicates assay cut-off.

**Supplemental Figure 3. Contribution of individual *SE* components to cross-reactivity suppression. (A)** DF patient samples La-18 S2, La-44 S2, La-90 S2, and La-99 S2 showing *SE*-mediated signal suppression in the DENV (NS1) IgG FcγR ELISA were retested in presence of individual *SE* components (recombinant flavivirus NS1 proteins). **(B)** Reactivity of samples La-18 S2, La-44 S2, La-90 S2, and La-99 S2 with HRP-labeled JEV NS1 in absence (w/o) and presence (w/) of the respective target-specific *SE* (comprising recombinant NS1 proteins from DENV, ZIKV, WNV, USUV, and YFV). *A*: absorbance, dotted line indicates assay cut-off, pos: positive.

