## Supplemental Material for "Nonstructural Protein 1 (NS1)-based IgG FcγR competition Enzyme-Linked Immunosorbent Assays (ELISAs) for highly specific serological profiling of previous flavivirus exposures"

### Supplemental Figure 1

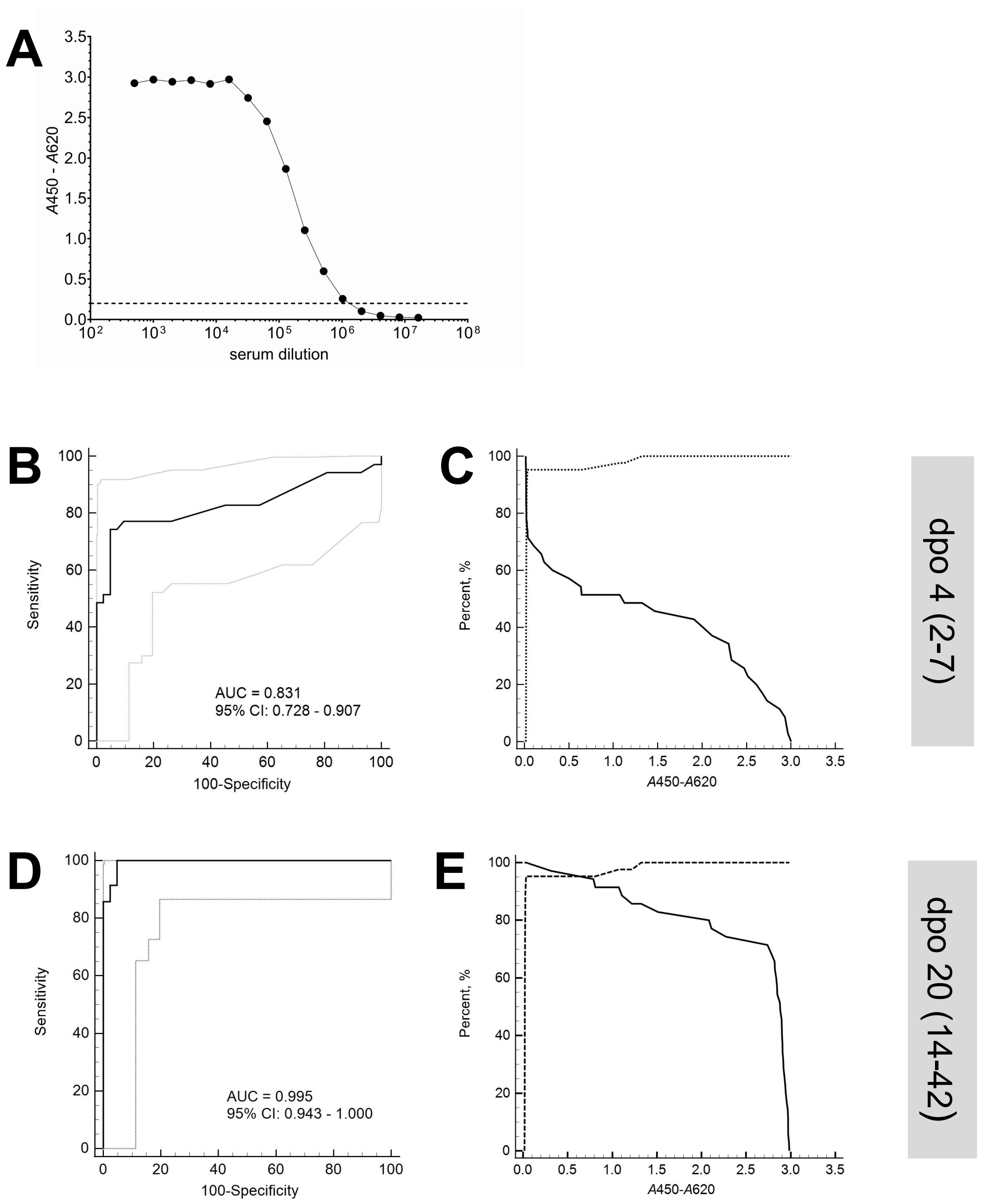


**Supplemental Figure 1. Detection range and ROC analyses (example DENV (NS1) IgG FcγR ELISA). (A)** Detection range. Serial dilutions of anti-DENV1-4 NS1 rabbit hyperimmune serum were analyzed with the DENV (NS1) IgG FcγR ELISA; dotted line indicates assay cut-off (*A*450 – *A*620 = 0.2). **(B) – (E)** ROC analysis was performed for paired serum samples from 35 Laotian patients with a PCR-confirmed DENV infection collected either between day 2 and 7 (median day 4) and between day 14 and 42 (median day 20) post onset of symptoms, respectively, and 42 serum samples from healthy European donors. *A*: absorbance, AUC: area under curve, CI: confidence interval.

### Supplemental Figure 2

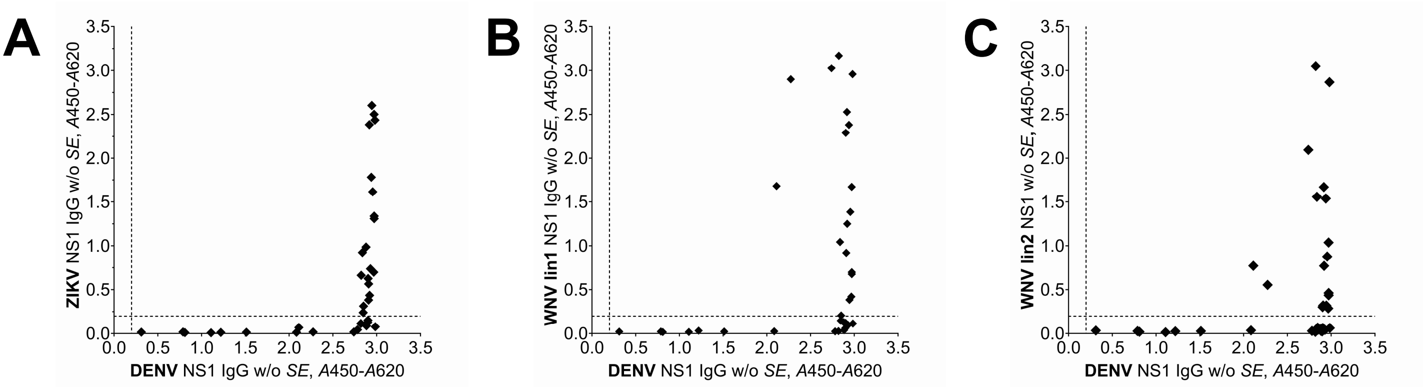


**Supplemental Figure 2. Correlation of DENV (NS1) IgG FcγR ELISA w/o *SE* results and (A) ZIKV, (B) WNV lin1, (C) WNV lin2 (NS1) IgG FcγR ELISA w/o *SE* results** for serum samples from 35 Laotian patients with a PCR-confirmed DENV infection, collected between day 14 and 42 post onset of symptoms. *A*: absorbance, dotted line indicates assay cut-off.

### Supplemental Figure 3

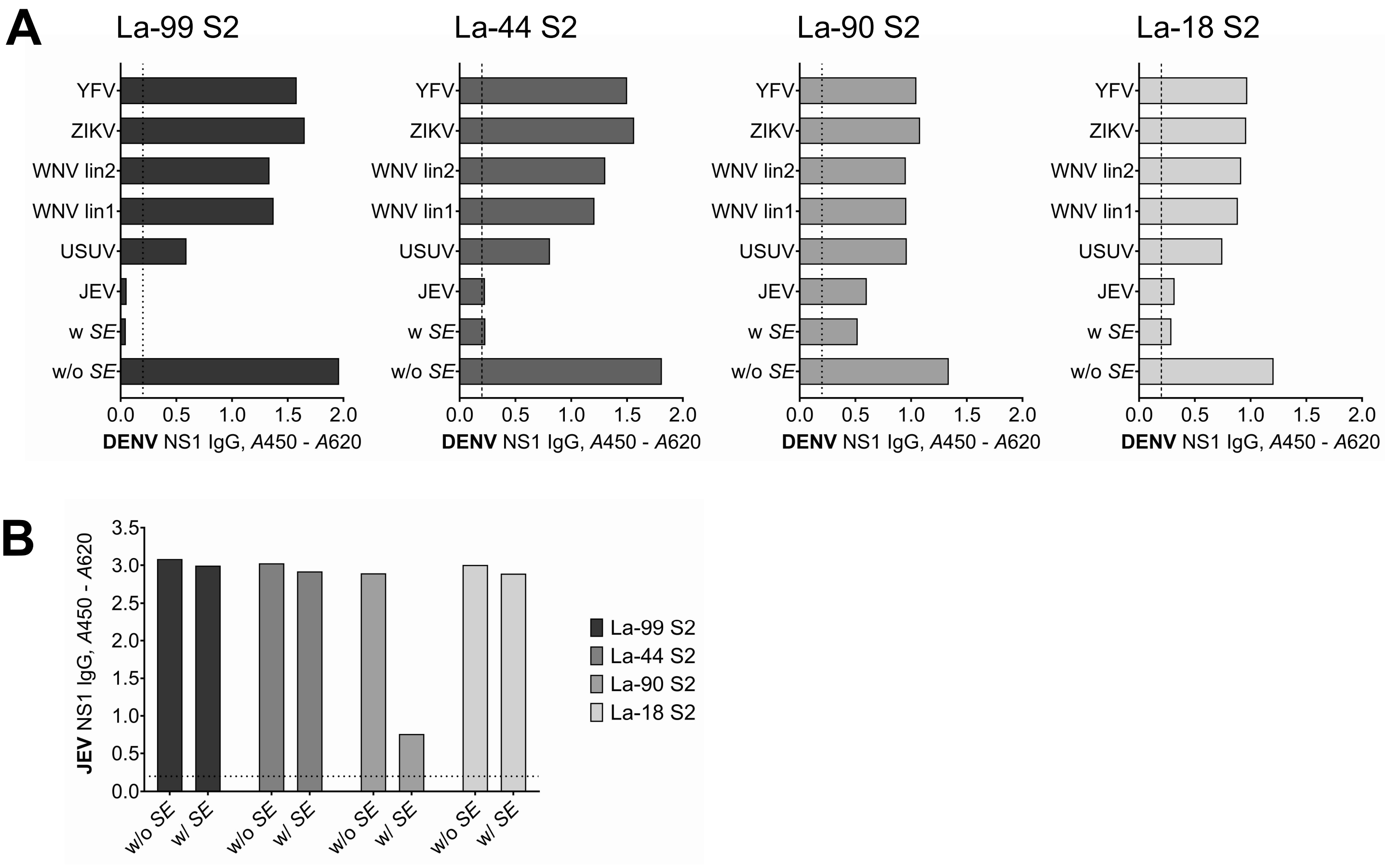


**Supplemental Figure 3. Contribution of individual *SE* components to cross-reactivity suppression. (A)** DF patient samples La-18 S2, La-44 S2, La-90 S2, and La-99 S2 showing *SE*-mediated signal suppression in the DENV (NS1) IgG Fc$\gamma$R ELISA were retested in presence of individual *SE* components (recombinant flavivirus NS1 proteins). **(B)** Reactivity of samples La-18 S2, La-44 S2, La-90 S2, and La-99 S2 with HRP-labeled JEV NS1 in absence (w/o) and presence (w/) of the respective target-specific *SE* (comprising recombinant NS1 proteins from DENV, ZIKV, WNV, USUV, and YFV). *A*: absorbance, dotted line indicates assay cut-off, pos: positive.

| group | subgroup | number | | sample information | provided by |
|---|---|---|---|---|---|
| **DENV infection** | German travelers | N = 12,<br>n = 37,<br>panel A | 2011 - 2017 | • residual samples from routine diagnostics<br>• PCR-confirmed DENV infection (3xDENV1, 6xDENV2, 1xDENV3, 2xDENV4)<br>• between 1 and 10 longitudinal samples per patient<br>• dpo: median 205 days, range 3 - 1224 days<br>• sex: 5 male, 5 female, 2 unknown<br>• age: median 35 years, range 18 - 63 years | BNITM, Hamburg, Germany |
| | | N = 11,<br>n = 35,<br>panel B | 2023 - 2024 | • residual samples from routine diagnostics<br>• PCR-confirmed DENV infection (4xDENV1, 2xDENV2, 4xDENV3, 1x unknown)<br>• between 1 and 6 longitudinal samples per patient<br>• dpo: median 9 days, range 2 - 1458 days<br>• sex: 5 male, 6 female<br>• 7 individuals with previous flavivirus vaccinations (JEV, TBEV, and/or YFV) | BNITM, Hamburg, Germany |
| | | N = 8 | 2011 - 2015 | • residual samples from routine diagnostics<br>• PCR-confirmed DENV infection (2xDENV1, 2xDENV2, 2xDENV3, 2xDENV4)<br>• sex: 3 male, 5 female<br>• age: median 55 years, range 25 - 71 years | BNITM, Hamburg, Germany |
| | Lao PDR residents | N = 35,<br>n = 70 | 2013 | • PCR-confirmed DENV infection (13xDENV2, 22xDENV3)<br>• one follow-up sample per patient<br>• dpo: S1 median 4 days (range 2 - 7 days), S2 median 20 days (range 14 - 42 days)<br>• sex: 17 male, 18 female<br>• age: median 20 years, range 15 - 55 years | BNITM/Savannakhet Provincial Hospital, Savannakhet, Lao PDR |
| | Colombia residents | N = 12<br>n = 24 | 2014 | • PCR-confirmed DENV infection (2x DENV1, 3x DENV2, 1x DENV3+4, 2x DENV4, 4x unknown)<br>• one follow-up sample per patient<br>• dpo: S1 median 4 days (range 3 - 8 days), S2 median 25 days (range 16 - 65 days)<br>• sex: 6 male, 6 female<br>• age: median 17 years, range 12 - 26 years<br>• 10 patients with previous YFV vaccination | BNITM/ Hospital Eduardo Arredondo Daza, Valledupar, Colombia |
| **DENV vaccination** | DENV naïve | N = 24,<br>n = 96 | 2023 - 2026 | • Qdenga vaccinees<br>• four longitudinal samples per vaccinee (S0: baseline, S1: ca. 30 days post first dose, S2: ca. 90 days post first dose, application of second dose, S3: ca. 120 days post first dose, ca. 30 days post second dose)<br>• sex: 11 male, 13 female<br>• age: median 30 years, range 22 - 67 years)<br>• 21 individuals reporting previous flavivirus vaccinations (TBEV, YFV, and/or JEV) | BNITM, Hamburg, Germany |
| | DENV preimmune | N = 25,<br>n = 100 | 2023 - 2026 | • Qdenga vaccinees<br>• self-reported previous DENV infection | BNITM, Hamburg, Germany |

| | | | | | |
|---|---|---|---|---|---|
| | | | | • four longitudinal samples per vaccinee (S0: baseline, S1: ca. 30 days post first dose, S2: ca. 90 days post first dose, application of second dose, S3: ca. days post first dose, ca. 30 days post second dose)<br>• sex:13 male, 12 female<br>• age: median 42 years, range 24 - 72<br>• 18 individuals with previous flavivirus vaccinations (TBEV, YFV, and/or JEV)<br>• years since reported DENV infection: median 1 year (no information from 4 participants) | |
| **ZIKV infection** | German travelers | N = 8,<br>n = 36 | 2015 - 2019 | • residual samples from routine diagnostics<br>• PCR-confirmed ZIKV infection<br>• several longitudinal samples per patient<br>• dpo: median 147 days, range 7 - 1240<br>• sex: 4 male, 4 female<br>• age: median 49 years, range 31 - 60 years<br>• 2 patients with previous flavivirus vaccinations (TBEV, YFV, and/or JEV) | BNITM, Hamburg, Germany |
| | | N = 9,<br>n = 14 | 2023 - 2025 | • residual samples from routine diagnostics<br>• 8 patients: ZIKV-PCR positive (urine), 1 patient: follow-up sample after PCR confirmation of ZIKV infection<br>• paired samples available for 5 patients (5 - 22 days between sampling time points) | BNITM, Hamburg, Germany |
| | Brazil residents | N = 9 | 2016 - 2017 | • PCR-confirmed ZIKV infection<br>• dpo: median 67 days (range 34 - 516 days)<br>• age: adults<br>• sex: 0 male, 9 female/pregnant<br>• ZIKV PRNT positive (titre >1280)<br>• positive in ZIKV (EDIII) IgG *SE* ELISA and Euroimmun ZIKV IgG ELISA | Fiocruz, Rio de Janeiro, Brazil |
| | | N = 5 | 2017 - 2018 | • dpo: median 1213 days (range 512 - 2178 days)<br>• age: children, median 3 years (range 1 - 6 years)<br>• ZIKV PRNT positive (titre range >160 - >640)<br>• positive in ZIKV (EDIII) IgG *SE* ELISA and Euroimmun ZIKV IgG ELISA | Fiocruz, Rio de Janeiro, Brazil |
| | WHO International Standard | N = 1 | not available | • 1st International Standard for anti-Asian lineage Zika virus antibody (human), NIBSC code: 16/352<br>• contains also antibodies reactive to DENV | NIBSC, South Mimms, UK |
| **WNV infection** | Ukraine residents | N = 8,<br>n = 12 | 2021 - 2022 | • residual samples from routine diagnostics<br>• WNV-IIFT IgM and IgG positive PE, ID<br>• dpo: median 15 days, range 4 - 32 days | Public Health Center of Ministry of Health of Ukraine, Kyiv, Ukraine |
| | Canada residents | N = 24 | not available | • WNV PRNT positive<br>• Focus WNV IgG DxSelect ELISA positive<br>• dpo: median 26 days, range 8 - 72 days | McMaster University, Hamilton, Canada |

| | | | | | |
|---|---|---|---|---|---|
| | Greece residents | N = 18 | 2024 - 2025 | • 4 samples from old infections (IgG avidity higher than 50%)<br>• 14 samples from acute infections (dpo 6 - 14), including 2 WNV PCR pos samples<br>• positive in Focus WNV IgG ELISA | Aristotle University of Thessaloniki, Thessaloniki, Greece |
| | Slovenia residents | N = 3,<br>n = 8 | 2018 - 2024 | • residual samples from routine diagnostics<br>• PCR confirmed WNV infection<br>• several longitudinal samples available from two patients | University of Ljubljana, Faculty of Medicine, Institute of Microbiology and Immunology, Ljubljana, Slovenia |
| | Germany residents | N = 3 | not available | • healthy blood donors<br>• WNV IgM/IgG ELISA positive | BNITM, Hamburg, Germany |
| **YFV vaccination** | German travelers | N = 91 | 2013 - 2014 | • sex: 38 male, 53 female<br>• age: median 31 years, range 18 - 82 | BNITM, Hamburg, Germany |
| **TBEV infection** | na | N = 2 | not available | • presumed TBEV infection (TBEV (NS1) IgG positive) | Biomex, Heidelberg, Germany; RKI, Berlin, Germany |
| | Slovenia residents | N = 15,<br>n = 45 | 2022 - 2023 | • residual samples from routine diagnostics<br>• longitudinal samples from confirmed TBEV infections (per patient: acute phase, 1-2 months post onset of symptoms, 1 year post infection)<br>• confirmed anti-TBEV NS1 IgG seroconversion | University of Ljubljana, Faculty of Medicine, Institute of Microbiology and Immunology, Ljubljana, Slovenia |
| ***Plasmodium* infection** | *P. falciparum* | N = 43 | 2012 - 2014 | • residual samples from routine diagnostics<br>• IIFT positive<br>• sex: 19 male, 8 female, 16 unknown<br>• age: median 41 years, range 13 - 75; 8 unknown | BNITM, Hamburg, Germany |
| | *P. malariae* | N = 11 | 2014 | • residual samples from routine diagnostics<br>• IIFT positive<br>• sex: 7 male, 3 female, 1 unknown<br>• age: median 41 years, range 18 - 58 | BNITM, Hamburg, Germany |
| **CHIKV infection** | na | N = 1 | 2016 | • WHO International Standard anti-CHIKV IgG (PEI code 1502/19)<br>• German patient who contracted chikungunya while traveling in Brazil in 2016<br>• pool of 3 donations 8 - 11 months after onset of symptoms<br>• negative for anti-DENV and anti-ZIKV antibodies | PEI, Langen, Germany |
| **SARS-CoV-2 infection** | Germany residents | N = 19 | 2020 - 2022 | • PCR-confirmed infections; strongly reactive in SARS-CoV-2 IgG IIFT and ELISA<br>• sex: 7 male, 12 female<br>• age: median 45 years, range 24 - 63 years | BNITM, Hamburg, Germany |
| **Healthy donors** | Germany residents | N = 42 | 2013 - 2020 | • unknown flavivirus immunstatus | BNITM, Hamburg, Germany |
| | Ukraine residents | N = 200 | before 2019 | • blood donor samples<br>• sex: 72 male, 128 female | |

| | | | | | |
|---|---|---|---|---|---|
| | | | | • 192 samples flavivirus IgG IIFT negative<br>• 8 samples flavivirus IgG IIFT positive | Public Health Center of Ministry of Health of Ukraine, Kyiv, Ukraine |
| | | N = 154 | 2021 | • blood donor samples | |
| | Colombia residents | N = 134 | 2014 | • sex: 55 male, 79 female<br>• age: median 27 years (range 17 - 51)<br>• 106 individuals with previous YFV vaccination | Hospital Eduardo Arredondo Daza, Valledupar, Colombia |
| | Lao PDR residents | N = 20 | 2014 | • sex: 3 male, 17 female<br>• age: median 48 years (range 23 - 53) | Savannakhet Provincial Hospital, Savannakhet, Lao PDR |
| | Ghana residents | N = 35 | 1998 - 1999 | • samples collected for onchocerciasis study | BNITM, Hamburg, Germany |

**Supplemental Table 1. Human serum samples;** N: number of patients/donors, n: number of samples. The table summarizes information of complete sample panels; subsets used for individual assay validations are stated in tables 2A-D; NIBSC: National Institute for Biological Standards and Control, BNITM: Bernhard Nocht Institute for Tropical Medicine, PEI: Paul Ehrlich Institute, RKI: Robert Koch Institute, dpo: days post onset of symptoms

| | | DENV (NS1) IgG | | ZIKV (NS1) IgG | | WNV lin1 (NS1) IgG | | WNV lin2 (NS1) IgG | | Presence of anti-flavivirus IgG confirmed by | | | | | |
|---|---|---|---|---|---|---|---|---|---|---|---|---|---|---|---|
| | | w/o *SE* | w/ *SE* | w/o *SE* | w/ *SE* | w/o *SE* | w/ *SE* | w/o *SE* | w/ *SE* | IgG IIFT | | | IgG ELISA | | |
| | **Sample ID** | *A*450 - *A*620 | *A*450 - *A*620 | *A*450 - *A*620 | *A*450 - *A*620 | *A*450 - *A*620 | *A*450 - *A*620 | *A*450 - *A*620 | *A*450 - *A*620 | DENV | ZIKV | WNV | Panbio DENV | Focus WNV | WNV (EP) FcγR |
| *Plasmodium malariae* | **1400 2556** | 0,018 | 0,013 | 0,020 | 0,020 | 0,040 | 0,029 | 0,025 | 0,025 | | | | | | |
| | **1400 6740** | 0,018 | 0,013 | 0,018 | 0,021 | 0,043 | 0,031 | 0,023 | 0,022 | | | | | | |
| | **1400 8974** | 0,019 | 0,015 | 0,021 | 0,023 | 0,048 | 0,033 | 0,033 | 0,034 | | | | | | |
| | **M019** | 0,018 | 0,017 | 0,020 | 0,019 | 0,031 | 0,030 | 0,021 | 0,021 | | | | | | |
| | **M020** | 0,025 | 0,025 | 0,026 | 0,024 | 0,084 | 0,073 | 0,045 | 0,046 | | | | | | |
| | **M021** | 0,016 | 0,014 | 0,020 | 0,021 | 0,044 | 0,042 | 0,029 | 0,029 | | | | | | |
| | **1400 6372** | 0,016 | 0,012 | 0,019 | 0,020 | 0,025 | 0,027 | 0,020 | 0,039 | | | | | | |
| | **1400 9297** | 0,019 | 0,014 | 0,020 | 0,019 | 0,045 | 0,042 | 0,026 | 0,029 | | | | | | |
| | **1401 0184** | 0,025 | 0,021 | 0,027 | 0,026 | 0,066 | 0,062 | 0,034 | 0,037 | | | | | | |
| | **1400 8902** | 0,021 | 0,017 | 0,023 | 0,020 | 0,037 | 0,034 | 0,025 | 0,030 | | | | | | |
| | **1400 9501** | 0,015 | 0,015 | 0,019 | 0,020 | 0,031 | 0,036 | 0,022 | 0,022 | | | | | | |
| *Plasmodium falciparum* | **1300 9308** | 0,017 | 0,011 | 0,021 | 0,017 | 0,034 | 0,024 | 0,019 | 0,024 | | | | | | |
| | **7476** | 0,014 | 0,011 | 0,017 | 0,018 | 0,040 | 0,036 | 0,018 | 0,020 | | | | | | |
| | **1300 5468** | 0,016 | 0,012 | 0,021 | 0,020 | 0,043 | 0,036 | 0,024 | 0,025 | | | | | | |
| | **1300 5508** | 0,018 | 0,013 | 0,021 | 0,020 | 0,033 | 0,028 | 0,020 | 0,022 | | | | | | |
| | **Pf64 (1300 1590)** | 0,020 | 0,015 | 0,021 | 0,023 | 0,050 | 0,036 | 0,030 | 0,035 | | | | | | |
| | **Pf65 (1300 5499)** | 0,112 | 0,074 | 0,020 | 0,020 | 0,049 | 0,034 | 0,031 | 0,026 | | | | | | |
| | **8143** | 0,017 | 0,015 | 0,020 | 0,019 | 0,052 | 0,038 | 0,023 | 0,025 | | | | | | |
| | **1300 4317** | 0,018 | 0,015 | 0,019 | 0,019 | 0,039 | 0,030 | 0,019 | 0,023 | | | | | | |
| | **1300 6992** | 0,015 | 0,013 | 0,019 | 0,018 | 0,059 | 0,030 | 0,018 | 0,021 | | | | | | |
| | **1300 7009** | 0,017 | 0,012 | 0,019 | 0,020 | 0,086 | 0,028 | 0,020 | 0,022 | | | | | | |
| | **1300 7256** | 0,019 | 0,014 | 0,020 | 0,019 | 0,116 | 0,062 | 0,022 | 0,026 | | | | | | |
| | **1300 7413** | 0,017 | 0,014 | 0,017 | 0,020 | 0,072 | 0,032 | 0,019 | 0,023 | | | | | | |
| | **1300 8505** | 0,014 | 0,014 | 0,018 | 0,017 | 0,029 | 0,030 | 0,017 | 0,023 | | | | | | |
| | **1300 6025** | 0,015 | 0,013 | 0,020 | 0,018 | 0,079 | 0,037 | 0,021 | 0,024 | | | | | | |
| | **1300 3822** | 0,016 | 0,010 | 0,020 | 0,021 | 0,042 | 0,031 | 0,030 | 0,022 | | | | | | |
| | **Pf57 (1301 3077)** | 0,018 | 0,013 | 0,022 | 0,019 | 0,044 | 0,046 | 0,021 | 0,022 | | | | | | |
| | **1301 0518** | 0,019 | 0,017 | 0,021 | 0,021 | 0,036 | 0,037 | 0,027 | 0,027 | | | | | | |
| | **Pf75 (1300 9751)** | 0,015 | 0,011 | 0,020 | 0,021 | 0,040 | 0,032 | 0,021 | 0,025 | | | | | | |
| | **1301 2902** | 0,017 | 0,013 | 0,022 | 0,021 | 0,040 | 0,028 | 0,020 | 0,020 | | | | | | |
| | **1300 5532** | 0,040 | 0,021 | 0,018 | 0,017 | 0,042 | 0,028 | 0,025 | 0,024 | | | | | | |
| | **1301 0059** | 0,022 | 0,016 | 0,022 | 0,022 | 0,049 | 0,038 | 0,037 | 0,032 | | | | | | |
| | **Pf67 (1300 9216)** | 0,015 | 0,010 | 0,018 | 0,019 | 0,033 | 0,024 | 0,023 | 0,020 | | | | | | |
| | **8931** | 0,123 | 0,062 | 0,020 | 0,020 | 0,042 | 0,036 | 0,030 | 0,030 | | | | | | |
| | **1300 5904** | 0,017 | 0,013 | 0,033 | 0,017 | 0,065 | 0,032 | 0,022 | 0,023 | | | | | | |
| | **1300 6224** | 0,015 | 0,012 | 0,019 | 0,019 | 0,072 | 0,044 | 0,020 | 0,023 | | | | | | |
| | **1300 9325** | 0,121 | 0,108 | 0,020 | 0,020 | 0,041 | 0,031 | 0,022 | 0,026 | | | | | | |
| | **1301 2524** | 0,016 | 0,013 | 0,018 | 0,019 | 0,031 | 0,028 | 0,019 | 0,023 | | | | | | |
| | **Pf2 (8559)** | 0,146 | 0,015 | 0,022 | 0,020 | 0,126 | 0,072 | 0,180 | 0,099 | | | | | | |
| | **Pf5 (9021)** | 0,074 | 0,060 | 0,020 | 0,017 | 0,044 | 0,029 | 0,025 | 0,028 | | | | | | |
| | **Pf66 (1300 2391)** | 0,108 | 0,104 | 0,019 | 0,019 | 0,042 | 0,033 | 0,020 | 0,019 | | | | | | |
| | **Pf68 (1300 9875)** | 0,033 | 0,024 | 0,024 | 0,021 | 0,105 | 0,071 | 0,051 | 0,050 | | | | | | |
| | **Pf69 (1300 9628)** | 0,019 | 0,015 | 0,022 | 0,020 | 0,049 | 0,036 | 0,024 | 0,024 | | | | | | |
| | **Pf20 (1300 5025)** | 1,154 | 1,065 | 0,025 | 0,018 | 0,037 | 0,034 | 0,026 | 0,025 | *not tested* | *not tested* | neg (< 1:20) | pos | *not tested* | pos |
| | **Pf56 (1300 2061)** | 0,392 | 0,336 | 0,025 | 0,020 | 0,046 | 0,032 | 0,027 | 0,028 | *not tested* | *not tested* | neg (< 1:20) | pos | *not tested* | pos |
| | **1300 5705** | 1,458 | 1,436 | 0,021 | 0,022 | 0,040 | 0,037 | 0,023 | 0,028 | pos (1:2560) | pos (1:320) | pos (1:2560) | pos | *not tested* | pos |
| | **1300 7215** | 1,581 | 1,369 | 0,020 | 0,018 | 0,099 | 0,035 | 0,020 | 0,022 | *not tested* | *not tested* | *not tested* | *not tested* | *not tested* | pos |
| | **Pf73 (1300 4144)** | 2,287 | 2,340 | 0,035 | 0,019 | 0,213 | 0,036 | 0,075 | 0,023 | pos (1:10240) | pos (1:320) | pos (1:2560) | *not tested* | pos | pos |
| | **1300 9268** | 0,287 | 0,261 | 0,050 | 0,037 | 0,236 | 0,113 | 0,210 | 0,107 | *not tested* | *not tested* | *not tested* | *not tested* | *not tested* | pos |
| | **Pf70 (9759)** | 1,686 | 0,062 | 0,039 | 0,020 | 2,352 | 0,574 | 2,886 | 1,497 | *not tested* | *not tested* | pos (>1:80) | *not tested* | pos | pos |
| | **Pf76 (9732)** | 0,605 | 0,012 | 0,019 | 0,020 | 1,147 | 0,276 | 0,817 | 0,197 | *not tested* | *not tested* | pos (>1:80) | *not tested* | pos | pos |
| | **Pf71 (1300 2698)** | 0,018 | 0,012 | 0,019 | 0,018 | 0,417 | 0,267 | 0,215 | 0,120 | *not tested* | *not tested* | pos (>1:80) | *not tested* | *not tested* | pos |
| | **Pf72 (1300 2557)** | 0,019 | 0,014 | 0,019 | 0,017 | 0,257 | 0,198 | 0,108 | 0,071 | *not tested* | *not tested* | pos (>1:80) | *not tested* | *not tested* | pos |
| | **Pf77 (1300 1815)** | 2,082 | 0,656 | 0,521 | 0,026 | 2,931 | 0,549 | 2,400 | 0,439 | pos (1:10240) | pos (1:2560) | pos (1:10240) | *not tested* | pos | pos |

**Supplemental Table 2. Analysis of *Plasmodium* IgG IIFT positive serum samples.** Samples (n=54) were tested with the DENV, ZIKV, and WNV (NS1) IgG FcγR ELISAs in absence (w/o) and presence (w/) of the assay-specific *SE* (cut-off *A*450 – *A*620 = 0.200, green: negative, orange: positive). Positive samples were subjected to confirmatory testing with flavivirus IgG IIFT and/or ELISA (Panbio DENV IgG ELISA, Focus WNV IgG ELISA, WNV Envelope Protein (EP) IgG FcγR ELISA (employing recombinant antigen comprising EP domains DI, DII, and DIII)).
